# ChatGPT sits the DFPH exam: large language model performance and potential to support public health learning

**DOI:** 10.1101/2023.07.04.23291894

**Authors:** Nathan P Davies, Robert Wilson, Madeleine S Winder, Simon J Tunster, Kathryn McVicar, Shivan T Thakrar, Joe Williams, Allan Reid

## Abstract

**Background:** Artificial intelligence-based large language models, like ChatGPT, have been rapidly assessed for both risks and potential in health-related assessment and learning. However, their application in public health professional exams have not yet been studied. We evaluated the performance of ChatGPT in part of the Faculty of Public Health’s Diplomat exam (DFPH).

**Methods:** ChatGPT was provided with a bank of 119 publicly available DFPH question parts from past papers. Its performance was assessed by two active DFPH examiners. The degree of insight and level of understanding apparently displayed by ChatGPT was also assessed.

**Results:** ChatGPT passed 3 of 4 papers, surpassing the current pass rate. It performed best on questions relating to research methods. Its answers had a high floor. Examiners identified ChatGPT answers with 73.6% accuracy and human answers with 28.6% accuracy. ChatGPT provided a mean of 3.6 unique insights per question and appeared to demonstrate a required level of learning on 71.4% of occasions.

**Conclusions:** Large language models have rapidly increasing potential as a learning tool in public health education. However, their factual fallibility and the difficulty of distinguishing their responses from that of humans pose potential threats to teaching and learning.

## Introduction

ChatGPT is an artificial intelligence (AI) chatbot that runs on OpenAI’s Generative Pre-Trained Transformer (GPT) models.^1^ It is one of a growing number of publicly available large language learning models (LLMs) that have been trained on huge volumes of text, using both machine learning and some human supervision, to help it respond to users in a conversational manner.

There have been concerns raised about the potential for LLMs to cause public health harm. This includes the possibility that LLMs like ChatGPT risk creating *infodemics* by generating vast amounts of plausible-sounding but incorrect information in both the research and public information spheres ^2^. Some, including the chief executives of major AI companies, warn that general artificial intelligence poses serious public health threats comparable to pandemics and nuclear war, as it has the potential for biological weaponisation, generate large-scale misinformation, and to strengthen the power of dictatorships.^3^ AI can be considered as a commercial determinant of health; a set of private sector activities which have a significant impact on health.^4^ As with other technologies,^5^ there may be a conflict between profit generation for AI companies and public health.

AI and LLMs have generated significant interest in health education. ChatGPT has performed relatively well on US medical^6,7^ and plastic surgery exams^8^ although it performed less well on the UK BioMedical Admissions Test^9^ and the Taiwanese Pharmacist Licensing Examination.^10^ Its novel abilities have generated discussions on its potential applications for medical teaching and learning.^13^

Public health exams often differ from biomedical exams. They are less likely to take multiple-choice or purely fact-based formats, requiring application of a broad range of concepts to open-ended scenarios. One such example is the Diplomate exam (DFPH), set by the Faculty of Public Health (FPH).^14^ Passing this exam is mandatory for progressing in public health specialty training in the United Kingdom. The DFPH exam is split into Paper 1 and Paper 2, sat sequentially. Paper 1 covers a broad range of topics, including research methods and epidemiology, screening, ethics, health promotion, health protection, sociology, leadership and management, health economics, health informatics, and healthcare public health.

We aimed to evaluate the performance of ChatGPT 3.5 in Paper 1 of the DFPH exam, including whether its answers were distinguishable from human respondents, and to investigate the level of insight and degree of learning it appeared to display.

## Methods

The 7 most recently available Paper 1s were selected from the Faculty of Public Health’s publicly available question bank (January 2014 – January 2017). Paper 1 incorporates 10 questions that require short, medium and long-form responses. It is divided into 5 topic-based sections, each with 2 questions. Papers from pre-2014 were excluded, as they comprise 10-mark essay-style questions. These differ significantly from the current style of questions, which are always broken down into at least two parts.

To generate responses from ChatGPT, each question component was entered, formatted by the question text followed by the direct question separated by a new line. For long-form answers, ChatGPT was given a prompt to write in full sentences rather than use bullet points. Responses were generated in February 2023 using ChatGPT version 3.5. Sessions were expunged after each question to avoid biasing.

Where the exam question required an answer “with regards to a particular country” or “with regards to a particular public health strategy”, the question was edited to be specific, for example “with regards to a public health obesity strategy”. This was to ensure the answer was specific to the countries and topics covered by the exam.

All 10 mark questions were excluded and all questions that include an image or require graphical output were also removed, as ChatGPT 3.5 was unable to parse images. Very light editing of the structure of the introduction to ChatGPT responses was required to maintain blinding, because ChatGPT often followed a very similar structure. American English was changed to British English. ChatGPT answers are provided in the supplementary material.

Questions were independently double-marked by two active DFPH examiners, using the DFPH exam moderation process to agree a final mark. These two examiners work as a pair in the real sittings of this exam. Prior to January 2017, candidates were required to score at least 50% in order to pass a question and could not fail more than two individual questions, so these were the criteria used to judge pass/fail.

Examiners were provided with a set of blinded answers for four papers with the lowest numbers of excluded questions; January 2017, June 2016, January 2016 and June 2014. 80% of answers were generated by ChatGPT and 20% of answers were from a bank of public health registrars preparing to sit the DFPH exam. Examiners were asked to indicate which answers they believed were generated by ChatGPT and which came from public health registrars.

Five public health registrars preparing for the DFPH exam, working in pairs, first independently measured the number of insights ChatGPT offered per answer for the full 7 exam papers, then came together to moderate scores. This used a modified definition of insight based on the work of Kung et al^7^, which must meet the following three criteria:

- Nondefinitional: Does not simply define a term in the input question
- Nonobvious: Requires deduction or knowledge external to the question input
- Valid: Is in keeping with public health practice or numerically accurate; preserves directionality

An example is provided in the supplementary material.

The same registrars then worked in pairs to judge each question against Bloom’s revised taxonomy of learning^15^ (BRT) assessing the level of learning ChatGPT appeared to be exhibiting in its answers against the level of learning those same registrars judged was required to answer the question appropriately. Training was provided to improve interrater reliability. Registrars assessed the level of learning required to answer the questions first before assessing the ChatGPT responses to avoid anchoring bias.^16^

## Results

### ChatGPT performance

Each of the 7 papers comprised of 10 questions worth 10 marks each, most of which were broken down into component parts. 21 out of 70 possible questions were removed (12 out of 40 of those marked). ChatGPT provided 119 individual responses across 7 exams. Results are provided in full in the supplementary material.

ChatGPT answers for whole questions scored between 4 - 9.5 out of a possible 10. Human answers ranged from 3.25 to 8.

ChatGPT averaged more than 5 out of 10 for each of four exams that were marked (Figure 1). However, it scored under 5 marks for 4 separate questions for the January 2017 paper, which would have resulted in failing the exam. ChatGPT would have been awarded a pass on 3 out of 4 exams. In comparison, recent pass rates for all of those who sat Paper 1 range from 47% to 65%.^14^ ChatGPT achieved a mean of 5.9 marks per question; the human respondents achieved a mean of 6.47.

**Figure 1.**
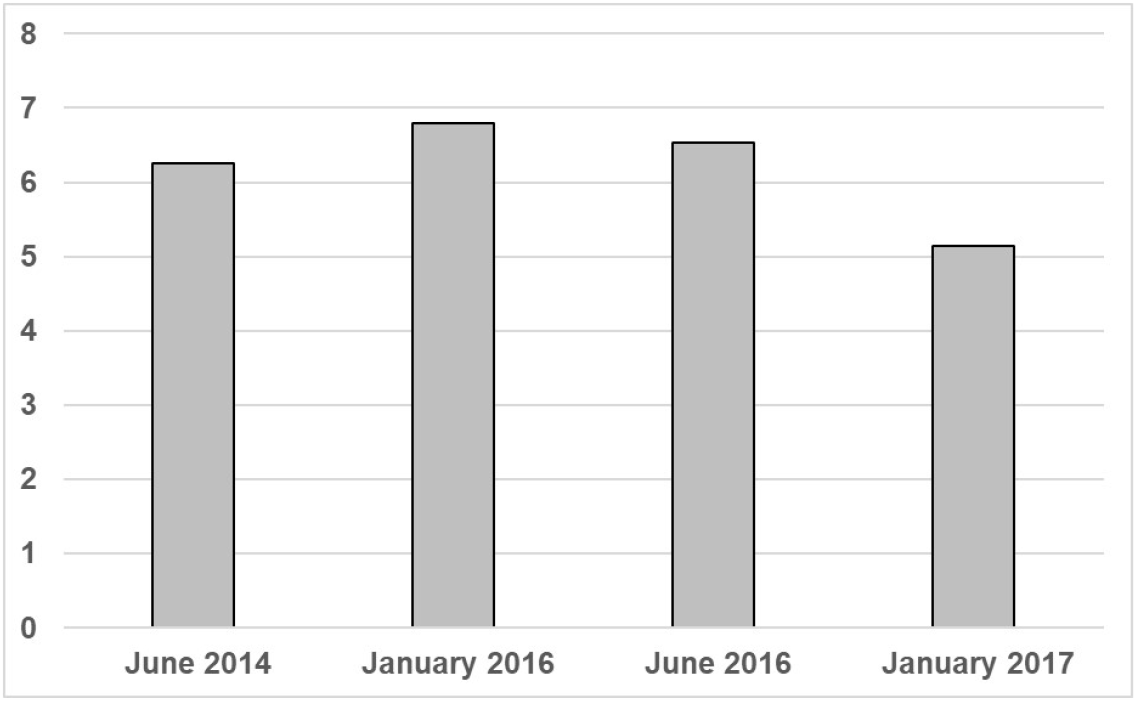
Mean ChatGPT score per exam

ChatGPT provided stronger responses on research methods than any other section, scoring an average mark of 7.95 in this question area. Its score in each of the other four sections were only just above a pass (Figure 2).

**Figure 2.**
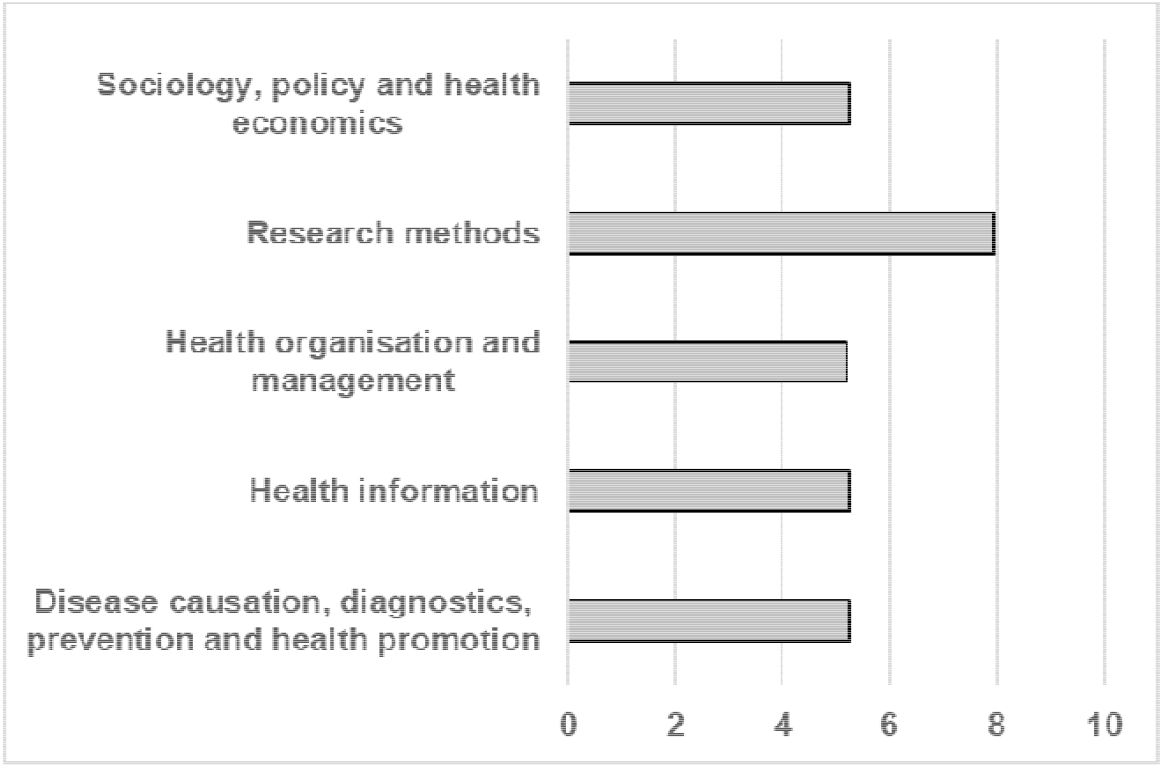
Mean ChatGPT mark per exam section

### Marker identification of respondent

Markers were able to identify that an answer was from ChatGPT in 39 of 54 instances (73.6% accuracy). However, they were only able to identify human answers in 4 out of 14 instances (28.6% accuracy).

### Unique insights

ChatGPT averaged 3.6 unique insights per question part. ChatGPT provided the greatest density of insight (around 4 per question part) for research methods, health information and health organization and management (Figure 3). The single score intraclass correlation for markers was 0.654 (95% CI 0.538 – 0.746).

**Figure 3.**
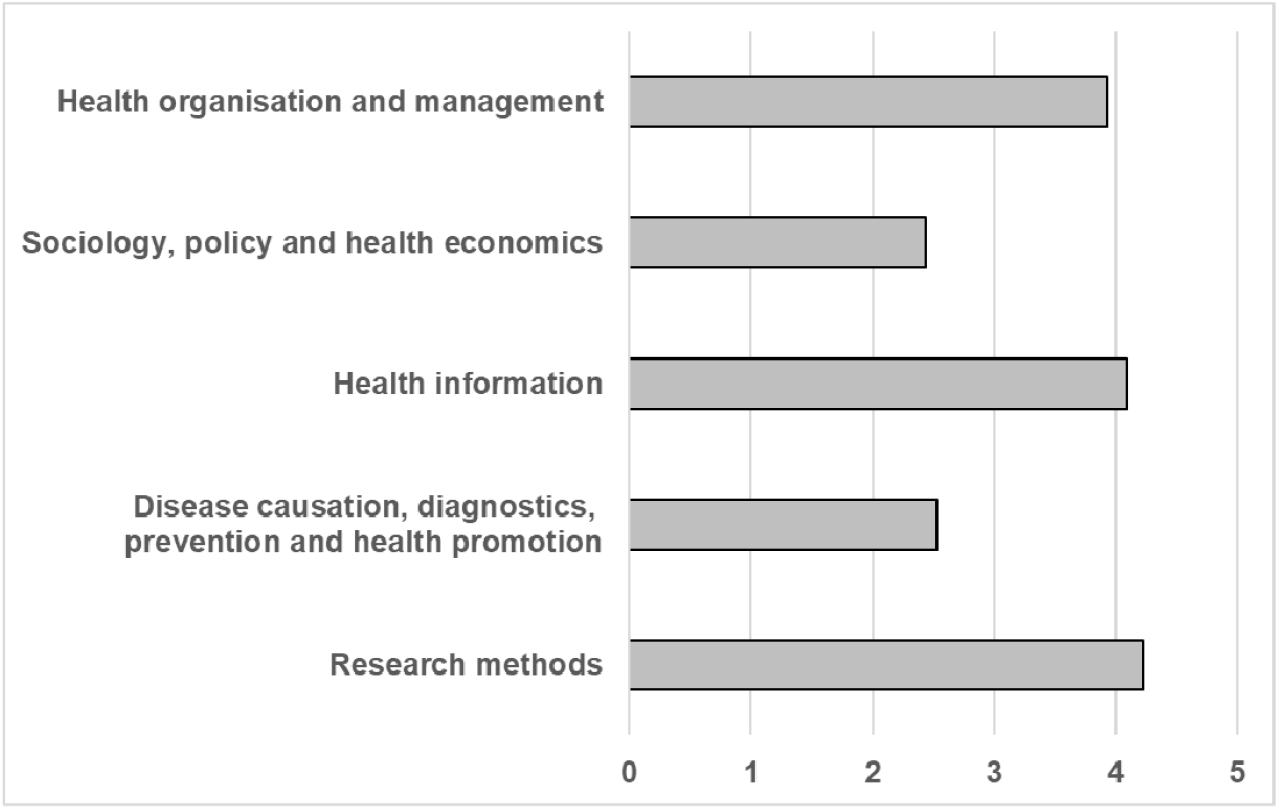
Mean ChatGPT density of insight per question part by section

### Bloom’s Revised Taxonomy (BRT)

71.4% of ChatGPT answers were judged to be at the ideal level on BRT and only 6.4% were two or more levels below (Figure 4).

**Figure 4.**
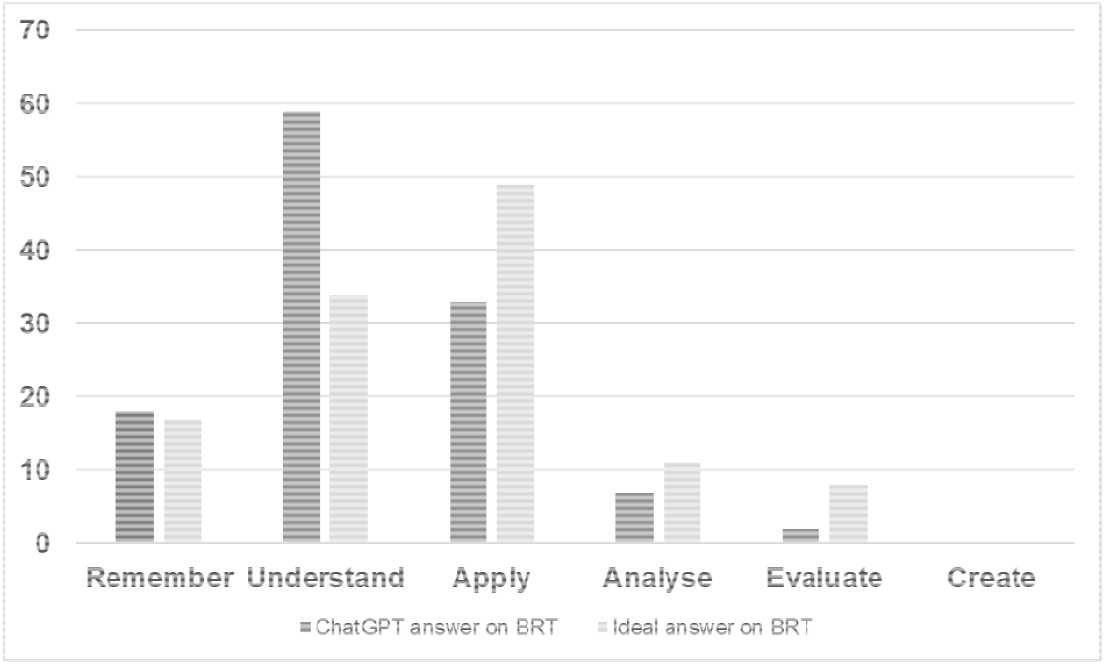
ChatGPT answer on BRT compared to ideal level

## Discussion

### Main findings of this study

We found that ChatGPT would have scored a pass mark in Paper 1 of the DFPH exam on 3 of 4 occasions. It had a higher floor to its answers than human respondents, never scoring below 4 marks, indicating that the textual corpus that it trained on enabled reasonable answers on the range of questions posed in DFPH Paper 1. Its scores per exam were very consistent, with all between 5 and 7. Much of the strength of its overall mark came from the research methods section, in which it scored an overall average of approximately 8, which is consistent with OpenAI’s findings that ChatGPT performs well in SAT Math and AP Statistics.^12^ It was very difficult for markers to differentiate between human answers and ChatGPT answers.

ChatGPT was able to generate non-obvious insights for each of the questions that it answered, which could be useful in supporting learning for students and those preparing for public health examinations. Its answers more often than not mimicked the requisite level of learning that a question required, which provides some evidence for its usefulness as a revision tool. For example, LLMs may be able to generate example questions that require a similar level of understanding to real public health exams for students to practice on.

However, it did provide inaccurate information, such as suggesting that deliberately infecting people with the bacteria that causes tuberculosis could form part of testing the efficacy of an intervention.

### What is already known on this topic

LLMs have the potential to support public health work in a number of areas, such as supporting coding and analysis, but also poses a series of threats, such as large-scale hallucination of information relating to public health, possible generation of bioweapons and potential strengthening of authoritarian regimes.

ChatGPT has variable performance in a range of health and biomedical examination scenarios. Some authors have suggested it could form a useful tool for revision and learning for students.

### What this study adds

This study shows that ChatGPT can generate plausible responses to a range of public health questions that were close to indistinguishable to answers from human public health registrars. The hallucination of facts (confidently expressing factually incorrect statements) remains an issue; whereas new versions of LLMs can provide references for their answers, the references themselves are often also hallucinated.^17^ It appears to give greater insight when considering more fact-based questions such as those on epidemiology and research methods; however, confident hallucination of facts is also likely to be a greater problem here.

There are implications for professional membership bodies and universities in marking public health exams and essays that may have been partially generated by LLMs, and in those supporting those undertaking public health qualifications to understand the strengths and limitations of AI chatbots in education.

### Limitations of this study

Due to marker availability, we were only able to appraise Paper 1 of the DFPH and were not able to assess Paper 2, which comprises critical appraisal and statistics papers. We also had to remove several questions incompatible with the new style of exam, reducing the pool of answers. Based on test outputs, it is likely that ChatGPT 3.5 would have particularly struggled with long-form critical appraisal questions as it consistently did not go into the detail required, despite specific prompting. It is possible ChatGPT was trained on answer banks similar to those provided by the DFPH.

We did not use follow-up prompts, which could have increased the relevance of answers further and supported review of use of ChatGPT as a learning aid. Although generating statistics on the density of insight for each question provides a broad overview of the usefulness of ChatGPT output, qualitative study into how LLMs work in practice as a revision tool is likely to be useful.

One limitation is that ChatGPT has already progressed to version 4.0, and independent medical researchers^11^ and OpenAI^12^ have both reported advancements over 4.0 on common assessment.^12^ Several other models, such as Google’s Bard, have also recently become available. Rapid assessment of each new iteration of LLMs in public health education would be required to keep abreast of its changing strengths and weaknesses.

Finally, this study very specifically examined ChatGPT performance in one particular exam. We must be wary of drawing broader conclusions on the use of AI in public health; this is a very specific scenario with lots of available material online. One area where markers noted that ChatGPT was weaker was on making its answers more specific to the scenario being posed, particularly in more open-ended questions, which likely limited its score in the non-research methods sections. Public health practice is very context-specific to the health needs of the communities being served.

### Conclusions

ChatGPT 3.5 performed relatively well on the DFPH Paper 1, particularly on the research methods sections. Its answers were difficult to distinguish from human answers and it may have utility for public health learning, although its propensity to hallucinate facts requires addressing for its full potential to be realised. More broadly, AI is largely developed and owned by private actors. Independent research and verification of its capabilities for good and for ill will be of utmost importance in the months and years to come.

## Supporting information

Supplementary material - ChatGPT output

Supplementary material - example of unique insight

## Data Availability

The data underlying this article are available in the article and in its online supplementary material.

https://osf.io/bpq4j/

