## Supplementary material - ChatGPT output for "ChatGPT sits the DFPH exam: large language model performance and potential to support public health learning"

**Supplementary material: ChatGPT 3.5 output for DFPH Paper 1s between January 2014 – January 2017**

[January 2014 questions and mark scheme](https://www.fph.org.uk/media/1216/jan-14-final-examiners-key-points-and-comments.pdf)

**Question 1**

a) One example is improved patient satisfaction. Measuring patient satisfaction can help determine the effectiveness of the support workers in providing care and support to patients. It can also help identify areas for improvement.

Increased adherence to treatment: Healthcare support workers can help patients better understand and follow their treatment plans, leading to improved health outcomes. Measuring adherence can help evaluate the impact of the support workers on patient health.

Reduced hospital admissions: Healthcare support workers can help manage patients' conditions more effectively, reducing the need for hospitalization. Monitoring hospital admissions can help assess the effectiveness of the support workers in reducing healthcare utilization.

Improved clinical outcomes: Healthcare support workers can help improve patient health outcomes such as reducing blood glucose levels in diabetes or improving lung function in COPD. Measuring clinical outcomes can provide a quantitative assessment of the impact of the support workers on patient health.

b) Three key features of a time series study design:

Longitudinal design: A time series study follows the same individuals over a period of time, allowing the investigation of changes in the outcome of interest over time.

Repeated measurements: In a time series study, the same outcome is measured multiple times, allowing for an assessment of the trend over time.

Control for confounding variables: A time series study design allows for the control of confounding variables that may impact the outcome of interest. This can be done by controlling for known confounding variables or by using statistical methods to account for their effects.

c) Three limitations of a time series study and ways to reduce them:

Temporal ambiguity: It can be difficult to determine the cause and effect relationship between the intervention and the outcome, as changes in the outcome may occur for reasons other than the intervention. This can be reduced by using a well-designed study with appropriate control groups.

Data collection challenges: Long-term data collection can be difficult, as patients may drop out or change their treatment plans. This can be reduced by using robust data collection methods and ensuring patient retention in the study.

Intervening events: Intervening events such as changes in treatment, disease progression, or unmeasured confounding variables can impact the outcome of interest. This can be reduced by controlling for known confounding variables and using statistical methods to account for unmeasured confounding variables.

**Question 2**

a) Possible explanations for the finding of a link between higher traffic density outside schools in England and low performance of primary school pupils could be due to several factors such as air pollution, noise pollution, reduced physical activity, and increased stress. Higher traffic density can lead to an increase in air pollution levels, which has been linked to respiratory and neurological problems and reduced cognitive function in children. Furthermore, the noise pollution associated with high traffic density can negatively impact children's ability to concentrate and learn. Additionally, children living in areas with higher traffic density may have less opportunity to engage in physical activity, which is important for their physical and mental health. Finally, increased stress due to the noise and danger associated with high traffic volume can negatively impact children's cognitive function and ability to learn.

b) To replicate the study locally, variables such as traffic density, air pollution, noise pollution, physical activity levels, stress levels, and academic performance would need to be measured. Traffic density can be measured by counting the number of vehicles passing by a school at a given time. Air pollution levels can be assessed through measures of particulate matter and nitrogen dioxide. The decibel level can be measured to assess the level of noise pollution in the area. Physical activity levels can be measured through self-reported surveys or objective measures such as accelerometers, while stress levels can be measured through self-reported surveys or physiological measures such as cortisol levels. Finally, academic performance can be measured through standardised tests or grades.

c) A good outcome measure for a study of this type should be relevant, reliable, and valid. It should be directly related to the research question and measure the link between traffic density and academic performance. The measure should produce consistent results when measured multiple times and should accurately reflect the construct it is intended to measure. Furthermore, it should have a clear and well-defined scoring system, and it should be easy to administer and interpret.

**Question 3**

a) Single gene disorders:

Genetic testing can be highly useful in cases of single gene disorders, where a mutation in a specific gene is responsible for causing the disease. Examples of single gene disorders include cystic fibrosis, sickle cell anaemia, and Huntington's disease. These tests can be used for a range of purposes, such as:

Diagnosis: Genetic testing can confirm a suspected diagnosis of a single gene disorder. This can be important for ensuring appropriate medical management and support for the individual and their family.

Carrier screening: Genetic testing can be used to identify individuals who are carriers of a single gene disorder, even if they do not show any symptoms themselves. This can be important for family planning purposes, as it can help couples understand their risk of having a child with the disorder.

Prenatal testing: For couples who are identified as carriers of a single gene disorder, genetic testing can be used during pregnancy to determine if the foetus has inherited the mutation. This information can help parents make informed decisions about their pregnancy.

Predictive testing: In some cases, genetic testing can be used to predict whether an individual who is at risk of a single gene disorder will develop the condition later in life. This can be important for early intervention and management of the condition.

b) Disorders influenced by multiple genes:

Genetic testing can also be useful in cases of disorders influenced by multiple genes, such as cancer. In these cases, genetic testing can be used for a range of purposes, including:

Risk assessment: Genetic testing can help determine an individual's risk of developing certain types of cancer. For example, testing for mutations in the BRCA1 and BRCA2 genes can help identify individuals who are at a higher risk of developing breast and ovarian cancer.

Diagnosis: In some cases, genetic testing can be used to confirm a diagnosis of cancer. For example, testing for specific genetic mutations in lung cancer can help determine the most effective treatment approach.

Treatment selection: Genetic testing can also help guide treatment selection in some cases. For example, testing for mutations in the EGFR gene can help determine whether certain targeted therapies will be effective in treating non-small cell lung cancer.

Familial screening: Genetic testing can be used to identify family members who may be at risk of developing cancer due to a genetic mutation. This can help these individuals make informed decisions about their healthcare and cancer screening.

Research: Genetic testing can also be used to further our understanding of the genetic basis of complex diseases like cancer. This can help drive the development of new treatments and prevention strategies.

**Question 4**

a) Epidemiology

Meningococcal meningitis is a bacterial infection caused by the bacterium Neisseria meningitidis. It can lead to meningitis (inflammation of the membranes surrounding the brain and spinal cord) or septicaemia (blood poisoning). Meningococcal meningitis is a serious and potentially life-threatening illness that can affect people of all ages, but it is most common in infants and young children. In England, the incidence of meningococcal meningitis has decreased in recent years, with approximately 1,500 to 2,000 cases reported each year.

Control

The control of meningococcal meningitis in England is primarily achieved through vaccination. The MenACWY vaccine is recommended for all adolescents aged 14-18 years and for first-year university students up to the age of 25. In addition, close contacts of cases of meningococcal meningitis may be given antibiotics to reduce their risk of developing the disease. Improved hygiene practices, such as frequent handwashing, can also help reduce the spread of the bacterium. Early recognition of symptoms and prompt treatment with antibiotics are also critical in controlling the spread of the disease.

b) Epidemiology

Measles is a highly contagious viral illness caused by the measles virus. It is spread through the air when an infected person coughs or sneezes, and it can cause a range of symptoms, including a high fever, runny nose, cough, conjunctivitis, and a rash. In England, prior to the introduction of a vaccine, measles was a common childhood illness with hundreds of thousands of cases each year. However, following the introduction of the MMR (measles, mumps, rubella) vaccine, the incidence of measles has declined significantly, with a few hundred cases reported each year.

Control

The control of measles in England is primarily achieved through vaccination. The MMR vaccine is recommended for all children, with two doses given at ages 12-15 months and 3-5 years. In addition, booster doses are recommended for individuals who are at increased risk of exposure to the virus, such as healthcare workers and travellers to endemic areas. Improved surveillance and reporting of cases, as well as public health campaigns to promote vaccine uptake, are also important in controlling the spread of measles. In the event of an outbreak, measures such as case identification and isolation, contact tracing, and quarantine may be implemented to prevent further spread of the virus.

**Question 5**

a)

One of the main purposes of population-based cancer registration is to monitor the burden of cancer in a population. Cancer registries collect data on the incidence and prevalence of cancer, including information on the type, location, and stage of cancer, as well as patient demographics. This information is used to track trends in cancer incidence and mortality over time, and to identify disparities in cancer outcomes across different population groups. For example, by tracking the incidence of specific types of cancer, public health officials can better understand the burden of cancer in their community and allocate resources appropriately.

Another purpose of population-based cancer registration is to inform public health planning and policy. Cancer registries provide critical data for public health planning and policy decision-making. The data collected can be used to identify areas with high cancer burden, to prioritise cancer control efforts, and to allocate resources for cancer prevention and treatment. For example, data from cancer registries can be used to inform the development of public health campaigns aimed at reducing the risk of certain cancers.

A third purpose of population-based cancer registration is to support cancer research. Cancer registries provide a valuable resource for cancer researchers, allowing them to study the epidemiology of cancer and to identify risk factors and potential targets for intervention. The data collected can also be used to evaluate the effectiveness of cancer control strategies. For example, researchers might use data from cancer registries to compare the incidence of certain cancers in different population groups, and to determine if there are disparities in cancer outcomes that need to be addressed.

Finally, population-based cancer registration can be used to improve cancer care. Cancer registries can be used to monitor the quality of cancer care, including information on treatment patterns and outcomes, and to identify opportunities for improvement. The data collected can also be used to evaluate the impact of new treatments and to inform the development of clinical guidelines. For example, data from cancer registries can be used to monitor the effectiveness of new treatments and to identify areas where further improvement is needed.

b) I will set out the main features of a population-based cancer registry.

Comprehensive data collection. A population-based cancer registry must collect comprehensive data on all cases of cancer in the population it serves. This includes information on the type, location, and stage of cancer, as well as patient demographics and treatment information.

Population coverage. A population-based cancer registry must have complete coverage of the population it serves. This means that all cases of cancer in the population must be captured and recorded, regardless of the setting in which they are diagnosed or treated.

Quality control. A population-based cancer registry must have strong quality control procedures in place to ensure the accuracy and completeness of the data collected. This includes procedures for verifying the accuracy of the data, as well as regular audits to identify and address any errors or discrepancies.

Data dissemination. A population-based cancer registry must make its data available to the public and to researchers in a timely and accessible manner. This may include publishing annual reports or providing access to the data through a publicly accessible database.

Confidentiality and privacy. A population-based cancer registry must have strong privacy and confidentiality protections in place to ensure that the personal information of patients is protected. This may include using secure methods for collecting and storing data, and ensuring that the data is only used for approved purposes.

Ongoing maintenance and updating. A population-based cancer registry must be regularly maintained and updated to ensure that it remains current and accurate. This includes updating the database with new cases of cancer, as well as regularly reviewing and verifying the data to ensure its accuracy.

**Question 6**

a) Assessing the need for falls prevention services in a local government administrative area requires gathering information from various sources. The main types of information sources that I would use include data and statistics, community engagement and feedback, expert opinions and best practices, and consultations with healthcare professionals and service providers.

Data and statistics from national surveys or local health records would provide insights into the prevalence and incidence of falls and related injuries in the area. For instance, the National Health Service (NHS) England publishes data on falls-related emergency hospital admissions, and this could help identify hotspots where falls are most common. However, relying solely on data may not reflect the true picture of falls in the community, as many falls go unreported or untreated.

Community engagement and feedback through focus groups or surveys would enable a better understanding of the experiences and perceptions of people who have had falls or are at risk of falling. This could also help to identify the specific needs and preferences of different demographic groups, such as older people or those with underlying health conditions. However, community engagement may not always be representative, and it can be challenging to reach out to all stakeholders, especially those who are socially isolated.

Expert opinions and best practices from academic literature or reports from government bodies or non-profit organizations would provide evidence-based guidance on what works best for falls prevention. This could include recommendations on the most effective interventions, such as exercise programmes or home modifications, and the cost-effectiveness of these interventions. However, expert opinions may not be applicable to every context, and there may be conflicting views on what works best.

Consultations with healthcare professionals and service providers would enable a better understanding of the current provision of falls prevention services in the area and their effectiveness. This could include feedback on the availability and accessibility of services, any gaps in provision, and the quality of care. However, the views of healthcare professionals may not always reflect the experiences of patients, and there may be biases in their perceptions.

In summary, a combination of these information sources would provide a more comprehensive and balanced assessment of the need for falls prevention services in a local government administrative area.

b) When recommending indicators for monitoring the impact of a new falls prevention service, there are several considerations to take into account. Firstly, the indicators should be measurable and feasible to collect, with clear definitions and methods of measurement. For instance, the number of falls-related hospital admissions, the proportion of patients who complete a falls prevention programme, or the number of home assessments conducted could be useful indicators to monitor.

Secondly, the indicators should be relevant and sensitive to the goals and objectives of the falls prevention service. For example, if the service aims to reduce the incidence of falls in older adults, then measuring the number of falls-related hospital admissions in this population would be more relevant than measuring falls in younger age groups.

Thirdly, the indicators should be responsive to change, with the ability to detect improvements or declines in the service over time. This would require setting baseline measures and regularly monitoring and reporting on progress towards the set goals. For instance, if the service aims to reduce falls in the community by 10% over a year, then monitoring the indicators at regular intervals could provide insights into whether the service is on track to achieving its goals.

Lastly, the indicators should be aligned with the wider policy and strategic objectives of the local government administrative area. For example, if there is a wider objective to improve health and wellbeing in the community, then the indicators could be framed in a way that links falls prevention to these broader goals.

In conclusion, selecting the right indicators for monitoring the impact of a new falls prevention service requires careful consideration of the indicators' measurability, relevance, responsiveness, and alignment with wider policy objectives.

**Question 7**

a) Patient and public involvement (PPI) in the process of policy development is a crucial aspect of healthcare. From a sociological perspective, PPI is viewed as a way of empowering patients and promoting democratic governance in the healthcare system. It involves giving patients a voice in the decision-making process and ensuring that their views and experiences are taken into account when developing policies.

PPI can be seen as a form of participatory democracy, where patients are active participants in the process of policy formation, rather than passive recipients of policies made by healthcare professionals or government officials. It also helps to address the power imbalances that exist in the healthcare system, where patients are often seen as passive recipients of care, rather than active partners in their own health and well-being.

PPI can also improve the quality of healthcare policies by incorporating the perspectives and experiences of patients. This helps to ensure that policies are informed by the real-life experiences and needs of patients, rather than being based solely on the views of healthcare professionals or government officials. PPI can also lead to more effective and efficient policies, as patients are likely to identify practical solutions to problems and offer insights into what works and what doesn't.

b) As the chair of this group, there are several steps I would take to ensure that patients are enabled to participate fully in the policy development process. Firstly, I would ensure that the group is inclusive and representative of the diverse patient populations that the healthcare organization serves. This could be achieved by reaching out to patient groups, communities, and organizations, as well as through advertising the group to the wider public.

Secondly, I would create a welcoming and supportive environment for patients to participate. This could involve providing accessible meeting spaces, as well as ensuring that the group's communication materials and processes are inclusive and accessible to all.

Thirdly, I would ensure that patients have the necessary resources and support to participate effectively in the group. This could include providing training and support to help patients understand the policy development process and how they can contribute effectively.

Finally, I would work to ensure that the views and experiences of patients are taken into account in the policy development process. This could involve creating opportunities for patients to give their views and experiences, as well as working with healthcare professionals and government officials to incorporate these perspectives into policy recommendations.

In summary, as the chair of the patient and public involvement group, my goal would be to create a supportive and inclusive environment that empowers patients to participate fully in the policy development process.

**Question 8**

a) Opportunity cost refers to the cost of a chosen alternative. It represents the value of the next best alternative that must be given up in order to pursue a certain action or decision. In other words, opportunity cost is the value of the benefits that are foregone as a result of making a particular choice.

b) Cost per quality-adjusted life year (QALY) is a measure of the cost-effectiveness of a healthcare intervention or programme. It represents the total cost of the intervention divided by the number of quality-adjusted life years (QALYs) gained as a result of that intervention. A QALY is a unit of measurement that takes into account both the quantity and quality of life gained as a result of a health intervention.

c) Opportunity cost and cost per QALY are important concepts in the rationalisation of services in a healthcare delivery system. They allow decision makers to compare the costs and benefits of different healthcare interventions, and to make informed decisions about which interventions to invest in and which to reduce or eliminate.

Opportunity cost can be used to evaluate the trade-offs between different healthcare interventions. For example, if a health delivery system invests in a new drug to treat a certain condition, it is important to consider the opportunity cost of that investment. This means considering the costs of the new drug, as well as the benefits that would have been gained if the resources had been invested in another intervention instead.

Cost per QALY can be used to determine the most cost-effective healthcare interventions. By considering both the cost and the benefits of different interventions, cost per QALY helps decision makers to identify which interventions are the most cost-effective in terms of improving health outcomes. For example, if two interventions have similar costs but one provides more QALYs than the other, the intervention with the higher QALYs would be considered the more cost-effective option.

In conclusion, the concepts of opportunity cost and cost per QALY can be useful tools for rationalising services in a healthcare delivery system. They help decision makers to consider both the costs and benefits of different interventions, and to make informed decisions about how best to allocate resources to improve health outcomes.

**Question 9**

a) Clinical governance refers to a systematic approach to maintaining and improving the quality of clinical care in a healthcare organization. It involves the development and implementation of policies, procedures, and systems to ensure that the care provided meets the highest standards and is consistent with the latest evidence-based practices. Clinical governance encompasses a wide range of activities, including clinical audit, risk management, and continuing professional development.

b) In a English general practice, the principles of good clinical governance can be used to improve the quality of health care in several ways. Firstly, it is important to establish clear systems for monitoring and evaluating the quality of care provided. This may involve regular audits of clinical records, as well as seeking feedback from patients and staff. Secondly, it is important to establish systems for managing risks and ensuring that any incidents or adverse events are promptly reported, investigated and acted upon.

In addition, it is crucial to ensure that all healthcare professionals working in the practice have the necessary skills, knowledge, and competencies to deliver high-quality care. This may involve providing regular training and development opportunities, as well as providing access to evidence-based clinical guidelines and best practice advice.

Finally, good clinical governance also involves creating an open, transparent and inclusive culture that supports learning, reflection and continuous improvement. This can be achieved by promoting regular communication and collaboration between all members of the practice team and encouraging feedback and suggestions from patients.

Overall, the principles of good clinical governance are essential for ensuring that the care provided in a general practice is of the highest quality, safe, effective and responsive to the needs of patients.

**Question 10**

a) Comparing and contrasting two models of organizing and funding health services at a population level, we can look at the UK National Health Service (NHS) and the US Private-Public mix system.

The UK National Health Service (NHS) is a publicly funded and publicly provided health service. It provides comprehensive and universal coverage to all citisens, regardless of their ability to pay. The NHS is funded primarily through general taxation, with additional contributions from National Insurance. The organization of the NHS is centralised, with the Department of Health and Social Care responsible for setting national policies and guidelines. Healthcare delivery is decentralised, with a mix of public and private providers.

The US Private-Public mix system, on the other hand, is a combination of public and private healthcare. The public sector includes programmes like Medicare for the elderly and disabled, and Medicaid for low-income individuals. The private sector, on the other hand, is composed of private health insurance plans and out-of-pocket payments by patients. The organization of the system is decentralised, with private insurance companies and healthcare providers setting policies and prices.

b) The UK National Health Service (NHS) has several advantages that positively impact the health and wellbeing of the population. Firstly, the NHS provides universal coverage to all citisens, meaning that access to comprehensive healthcare services is not restricted based on the ability to pay. This results in a more equitable distribution of healthcare services and reduces the burden on individuals.

Another advantage of the NHS is its cost-effectiveness. It is funded primarily through general taxation, which helps to spread the costs of healthcare across the population. This reduces the burden on individuals, as they do not have to pay for health insurance, co-payments, or deductibles. Additionally, the centralization of the NHS allows for efficient planning and coordination of healthcare services, reducing waste and ensuring consistent standards of care.

However, the NHS also has its disadvantages. One major disadvantage is long waiting times for non-emergency procedures, as the high demand for services can result in patients having to wait for extended periods of time. Another disadvantage is limited choice of healthcare providers. Patients are restricted to the healthcare providers within the NHS system, and have little room for choice. Furthermore, the NHS is funded by general taxation, which can result in budget constraints. This can limit the amount of resources available for healthcare services, and may impact the quality of care.

In conclusion, the UK National Health Service (NHS) has several advantages and disadvantages that impact the health and wellbeing of the population. While it provides universal coverage and cost-effectiveness, long waiting times and limited choice of healthcare providers are also major challenges that the NHS faces.

The US private-public mix healthcare system has several advantages that impact the health and wellbeing of the population. Firstly, the combination of public and private healthcare provides individuals with more choice and control over their healthcare. Patients can choose from a range of private health insurance plans and out-of-pocket payments, allowing them to select the plan that best suits their needs and budget.

Another advantage of the private-public mix system is that it provides incentives for innovation and cost-effectiveness. Private healthcare providers and insurance companies are motivated to provide high-quality, cost-effective care in order to attract patients and maintain market share. This can result in the development and implementation of new and more effective treatments, as well as the introduction of cost-saving measures.

However, the US private-public mix healthcare system also has its disadvantages. One major disadvantage is the unequal distribution of healthcare services, as access to healthcare is largely based on the ability to pay. Those without insurance or without the means to pay out of pocket face significant barriers to accessing necessary care. Another disadvantage is the high cost of healthcare, as the private sector often sets prices and policies that prioritise profit over patient needs. This results in high costs for patients, including premiums, co-payments, and deductibles.

In conclusion, the US private-public mix healthcare system has several advantages and disadvantages that impact the health and wellbeing of the population. While it provides more choice and incentives for innovation, unequal distribution of care and high costs for patients are also major challenges that the system faces.

[June 2014 exam questions and mark scheme](https://www.fph.org.uk/media/1217/june-2014-part-a-questions-key-points-and-examiners-comments.pdf)

Question 1 – removed as graph-based content

Question 2

a) A case-control study could be an appropriate research design to investigate the hypothesis that sarcoidosis is caused by recent infection with a specific mycobacterium. In a case-control study, cases (people with sarcoidosis) are compared to controls (people without sarcoidosis) to see if there are any differences in their exposure to the potential risk factor, in this case, recent infection with the mycobacterium. This design is appropriate because it can quickly identify if there is an association between the risk factor and the disease.

The study would involve recruiting cases and controls from the same population and asking them about their exposure to the mycobacterium. Information about their medical history, lifestyle, and other potential risk factors would also be collected. The cases and controls would then be compared to see if there is a higher prevalence of recent infection with the mycobacterium among the cases compared to the controls. If there is a significant difference, it would provide evidence to support the hypothesis that recent infection with the mycobacterium is a cause of sarcoidosis.

b) Confounding variables are variables that are related to both the exposure and the outcome, making it difficult to determine if the exposure is actually causing the outcome. Confounding can result in an overestimate or underestimate of the true effect of the exposure on the outcome. Some general features of confounding variables include:

1. They are associated with the exposure and the outcome.
2. They are not intermediate variables in the causal pathway between the exposure and the outcome.
3. They are not a result of the exposure.
4. They are not the same as the exposure or outcome.

c) There are several techniques that can be used to reduce confounding in a study:

1. Matching: This involves selecting controls who are similar to the cases in terms of important confounding variables. For example, in a case-control study of sarcoidosis and mycobacterium infection, the cases and controls could be matched on age, sex, and other relevant factors.
2. Stratification: This involves dividing the study population into subgroups based on the confounding variables and then analysing each subgroup separately. For example, the study population could be divided into age groups and then the association between mycobacterium infection and sarcoidosis could be assessed in each age group.
3. Multivariable analysis: This involves adjusting for the confounding variables in a statistical model. For example, a regression model could be used to adjust for age, sex, and other confounding variables and then the association between mycobacterium infection and sarcoidosis could be assessed.
4. Randomization: This involves assigning the exposure randomly to participants in a study. For example, in a randomized controlled trial, participants could be randomly assigned to receive either the mycobacterium or a placebo, and then the incidence of sarcoidosis could be compared between the two groups. This would reduce the impact of confounding because the exposure would be assigned randomly and not based on the confounding variables.Top of Form

Question 3 – removed as 10 mark question

Question 4 – removed as 10 mark question

Question 5 – removed as graph-based content

Question 6

a) Surveys are widely used as a method of collecting health information, and they offer several advantages as well as some disadvantages.

Advantages:

Surveys are cost-effective and efficient methods of collecting data on a large scale. For example, in England, the Health Survey for England (HSE) is a yearly survey that collects data on the health of the population. It is designed to provide a representative sample of the population and is an efficient way of collecting data on a large scale.

Surveys can collect data on a wide range of health issues, including physical health, mental health, and lifestyle factors. For example, the National Child Measurement Programme (NCMP) in England uses surveys to collect data on childhood obesity rates and trends. This data is used to inform public health policy and interventions.

Surveys can be tailored to specific populations or health issues, allowing for targeted data collection. For example, the Health and Wellbeing Survey for London is a survey that collects data on the health and wellbeing of London residents, including data on specific health issues such as air pollution and access to green spaces.

Disadvantages:

Surveys rely on self-reported data, which can be subject to bias and inaccuracies. For example, individuals may underreport or overreport their health behaviours or symptoms. This can lead to inaccurate data and flawed conclusions.

Surveys can be time-consuming and resource-intensive. For example, the National Diabetes Audit in England is a yearly survey that collects data on diabetes care and outcomes. It requires a significant investment of time and resources to administer and analyse the data.

Surveys may not provide a complete picture of health issues, as they are limited to the questions asked. For example, a survey on mental health may not capture all mental health issues or provide a complete understanding of the social determinants of mental health.

b) Surveys are particularly useful for investigating aspects of health that are difficult to measure using routine sources. Some examples include:

1. Health behaviours and lifestyle factors: Surveys are useful for collecting data on health behaviours and lifestyle factors, such as diet, physical activity, and smoking status. For example, the Health Survey for England collects data on these factors, which are important determinants of health outcomes.
2. Patient-reported outcomes: Surveys are useful for collecting patient-reported outcomes, such as quality of life, symptoms, and satisfaction with care. These outcomes are important indicators of the effectiveness of healthcare interventions. For example, the National Cancer Patient Experience Survey in England collects data on cancer patients' experiences with care and treatment.
3. Rare or stigmatized health issues: Surveys are useful for collecting data on rare or stigmatized health issues, as routine sources may not capture this information. For example, the Gay Men's Sex Survey in England collects data on the sexual health and behaviours of gay and bisexual men, a population that may be underrepresented in routine health data.

Question 7

(a) Marginal analysis is a decision-making tool that involves comparing the additional benefits of an action with its additional costs. In other words, it involves weighing the costs and benefits of taking a specific action, such as making an investment, expanding production, or launching a new product, in order to determine whether it makes economic sense. Marginal analysis helps individuals and organizations make informed decisions by considering the incremental impact of each additional unit of an action on the costs and benefits involved.

(b) To illustrate how marginal analysis can be applied to a public health service or a hospital service in England, consider the example of a hospital considering whether to invest in a new medical imaging technology, such as an MRI machine. The hospital can use marginal analysis to determine whether the benefits of acquiring this technology, such as improved diagnostic accuracy and efficiency, outweigh the costs of purchasing, installing, and maintaining the machine.

For example, the hospital can calculate the marginal cost of acquiring and operating the MRI machine and compare it to the marginal benefits in terms of increased patient volume and improved diagnostic accuracy. If the marginal benefits exceed the marginal costs, then the hospital can proceed with the investment.

(c) Supply and demand analysis is a tool used to understand the behaviour of buyers and sellers in a market, and to predict changes in market conditions based on changes in demand and supply. It is based on the economic principle that prices are determined by the interaction of supply and demand in a market.

Supply refers to the quantity of a good or service that a producer is willing and able to offer for sale at a given price, while demand refers to the quantity of a good or service that a consumer is willing and able to buy at a given price. Market forces, such as changes in consumer preferences or production costs, can cause changes in supply and demand and lead to changes in market prices.

(d) To illustrate how the techniques of supply and demand analysis can be applied to a public health service or a hospital service in England, consider the example of a hospital that is experiencing a shortage of nurses. The hospital can use supply and demand analysis to understand the factors that are contributing to the shortage and to identify potential solutions.

For example, the hospital can analyse the supply of nurses by examining factors such as the number of qualified candidates available, the level of salaries offered, and the overall work environment. On the demand side, the hospital can examine factors such as patient volume, the complexity of the cases being treated, and the resources available for nurse training and development.

Based on this analysis, the hospital can develop strategies to address the shortage, such as offering higher salaries, improving the work environment, or investing in nurse training programs. By applying the principles of supply and demand analysis, the hospital can make informed decisions about how to improve its nurse staffing levels and provide better patient care.

Question 8

(a) The effects of unemployment on ill health can be significant and long-lasting. Unemployment has been linked to a range of physical and mental health problems. One of the most common effects of unemployment is increased stress and anxiety. This is because unemployment often leads to financial insecurity and a decreased quality of life, both of which can have a negative impact on a person's mental health.

In addition to increased stress and anxiety, unemployment has been associated with other mental health problems, such as depression, substance abuse, and suicide. Unemployment can also have physical health effects, such as increased rates of cardiovascular disease and reduced access to health care.

b) There are several possible social explanations for the relationship between unemployment and health. One of the most common explanations is the stress and anxiety associated with unemployment. As previously mentioned, unemployment can lead to financial insecurity and decreased quality of life, which can cause increased stress and anxiety.

Another possible explanation is the loss of social support and community that often occurs with unemployment. For many people, work provides a sense of purpose and social connection, and losing this can have a negative impact on mental and physical health.

Additionally, unemployment can lead to a reduction in access to health care, as many people lose their health insurance along with their jobs. This can result in delayed or inadequate treatment for health problems, which can in turn lead to more serious health problems and increased health care costs in the long run.

Overall, the relationship between unemployment and health is complex, and is likely influenced by a variety of social, economic, and psychological factors.

Question 9

(a) Three approaches that could be used to assess whether levels of healthcare spending by an organization need to be changed are:

Cost-Benefit Analysis: This approach involves evaluating the costs and benefits of healthcare spending and determining whether the benefits outweigh the costs. This approach is often used to make decisions about the allocation of limited resources.

Cost-Effectiveness Analysis: This approach evaluates the costs and effectiveness of healthcare spending, comparing different programs or interventions in terms of the costs and health outcomes achieved. This approach is useful for identifying areas where spending can be reduced or reallocated to achieve better health outcomes.

Budget Impact Analysis: This approach evaluates the financial implications of changing healthcare spending levels, including the impact on revenue and expenses. This approach can be useful for organizations that need to balance the need to improve health outcomes with the need to manage costs.

(b) Cost-Benefit Analysis

Cost-Benefit Analysis is an approach that assesses the costs and benefits of a particular intervention or program in order to determine whether the benefits outweigh the costs. In the context of healthcare, this approach can be used to evaluate whether increasing or decreasing spending on a particular programme or service will result in a net positive impact on health outcomes.

The strengths of Cost-Benefit Analysis include its focus on the financial implications of healthcare spending, which can be important for organizations that need to balance the need to improve health outcomes with the need to manage costs. Additionally, this approach can provide a clear and objective comparison of the costs and benefits of different programs or interventions.

The weaknesses of Cost-Benefit Analysis include the difficulty of quantifying certain benefits, such as improved health outcomes, and the difficulty of predicting the future costs of a particular intervention or program. Additionally, this approach can sometimes ignore other important considerations, such as ethical or moral considerations, that are not easily quantifiable in financial terms.

Overall, Cost-Benefit Analysis is a useful approach for organizations that need to make decisions about healthcare spending, but it should be used in conjunction with other approaches, such as Cost-Effectiveness Analysis and Budget Impact Analysis, to ensure that all relevant factors are taken into account.

Question 10

(a) Conflict can occur within a project team in several circumstances. One of the most common causes of conflict is differences in opinions or perspectives among team members. For example, team members may have different ideas about the best approach to solving a particular health issue, which can lead to disagreements and conflicts.

Another cause of conflict within a project team is competition for resources. When resources are limited, such as funding, equipment, or manpower, team members may compete for these resources, leading to conflicts.

Additionally, conflicts can arise when team members have different goals and priorities. For example, one team member may prioritise research, while another may prioritise providing care to patients. When these goals and priorities clash, conflict can result.

(b) To resolve conflicts within a project team, several techniques can be used. One effective technique is active listening and communication. This involves encouraging all team members to express their opinions and concerns and to listen actively to one another. By creating an environment where all team members feel heard and understood, conflicts can often be resolved more effectively.

Another effective technique is mediation or facilitation. This involves bringing in an impartial third party, such as a mediator or facilitator, to help the team work through the conflict. The mediator or facilitator can help team members understand each other's perspectives, identify common ground, and find mutually acceptable solutions.

A third technique is collaboration and negotiation. This involves encouraging team members to work together to find a solution that meets everyone's needs and concerns. By working together, team members can often find creative and mutually acceptable solutions to conflicts.

Finally, it is important for team leaders to promote a positive and constructive work environment. This includes setting clear goals and expectations, promoting open communication, and addressing conflicts promptly and effectively. By creating a positive and supportive work environment, conflicts within the team can be reduced, and when conflicts do arise, they can be more easily resolved.

[January 2015 questions and mark scheme](https://www.fph.org.uk/media/1214/january-15-key-points-examiners-and-chair-comments.pdf)

**Question 1**

a)

Cluster randomisation refers to the process of randomly allocating whole groups of people, rather than individuals, to either an intervention or control group. For example, in a study to evaluate the effectiveness of a new school-based nutrition programme, schools could be randomly allocated to either receive the intervention or act as the control group. All students within a school would then be included in the same group, rather than randomly selecting individual students to be included in each group.

b)

A crossover design is a type of experimental design in which each participant receives both the intervention and control treatments, but in a different order. For example, in a study to evaluate the effectiveness of a new drug for treating headaches, each participant could receive the drug for a period of time followed by a placebo for a similar period of time, or vice versa. The aim of a crossover design is to control for individual differences and increase the efficiency of the study.

c)

Intention to treat is a principle in clinical trials that states that all participants should be analysed in the group to which they were originally assigned, regardless of whether or not they completed the intervention or adhered to the protocol. For example, in a study to evaluate the effectiveness of a new diet for weight loss, participants who were originally assigned to the intervention group but did not complete the programme should still be included in the analysis of the intervention group, rather than being excluded. The intention to treat principle helps to reduce the risk of bias in the results of a study.

**Question 2**

a) Strategies for sampling the population:

Simple Random Sampling. In this method, each member of the population has an equal chance of being selected to participate in the survey. This is a straightforward and unbiased approach, but it can be time-consuming and impractical when the population is large.

Stratified Random Sampling: In this method, the population is divided into subgroups (strata) based on certain characteristics (e.g. age, gender, location), and a random sample is selected from each stratum. This approach helps to ensure that the sample is representative of the population and allows for comparisons between different subgroups. However, it can be more complex to implement and requires a good understanding of the population.

Cluster Sampling. In this method, the population is divided into groups (clusters), and a random sample of clusters is selected. All members of the selected clusters are then included in the survey. This approach is less precise than simple or stratified random sampling, but it is often more practical and cost-effective, especially when the population is spread out over a large geographic area.

b) Types of questions to include in the questionnaire:

Demographic questions. These questions aim to gather information about the background and characteristics of the participants (e.g. age, gender, education level, occupation, etc.).

Attitudinal questions. These questions aim to assess the attitudes and opinions of the participants towards the park facilities (e.g. "How satisfied are you with the quality of the park facilities?").

Behavioural questions. These questions aim to gather information about the participants' actual behaviours and usage patterns (e.g. "How often do you visit the park?").

Satisfaction questions. These questions aim to assess the level of satisfaction with different aspects of the park facilities (e.g. "How satisfied are you with the cleanliness of the park?").

Open-ended questions. These questions allow participants to provide their own answers in their own words (e.g. "What do you think could be improved about the park facilities?").

c) Considerations for questionnaire design:

The number of questions - The number of questions should be kept to a minimum to avoid participant fatigue and to increase the response rate.

The order of questions - The order of questions should be carefully considered to ensure that the questionnaire flows logically and is easy to follow. Demographic questions, for example, could be placed at the beginning, while more sensitive or personal questions could be placed towards the end of the questionnaire.

Question wording - The wording of questions should be clear, concise, and neutral to avoid leading the participants to a particular answer.

Question format - The format of questions (e.g. multiple choice, Likert scale, open-ended) should be appropriate for the type of information being gathered and should be easy for the participants to understand and respond to.

Question 3 – removed as 10 mark question

Question 4 – removed as 10 mark question

Question 5

a) Potential sources of routinely collected administrative data in England that could be used to replace a periodic census include:

National Health Service (NHS) records. This database contains information on the demographics, health status, and treatment history of individuals in England, making it a valuable source of information for population health research and analysis.

Department for Work and Pensions (DWP) records. This database contains information on the employment and benefits history of individuals in England, providing valuable information on the economic status of the population.

Department for Education (DfE) records. This database contains information on the educational history and attainment of individuals in England, providing valuable information on the educational status of the population.

Electoral Register. This database contains information on the demographic characteristics and voting history of individuals in England, providing valuable information on the demographic makeup and political engagement of the population.

National Insurance number (NINo) records. This database contains information on the demographic characteristics and employment history of individuals in England, providing valuable information on the employment status and demographic makeup of the population.

b) Advantages of routinely collected administrative data for generating population level statistics:

1. Cost: Administrative data is collected for other purposes and is often available at a lower cost than conducting a census, making it a more cost-effective option for generating population level statistics.
2. Timeliness: Administrative data is collected on an ongoing basis, providing up-to-date information on the population that can be used for population level analysis.
3. Comprehensive coverage: Administrative data often covers a large and diverse portion of the population, making it a valuable source of information for population level analysis.
4. Reduced response burden: Using administrative data reduces the response burden on individuals, as they do not need to participate in a census or survey.

Disadvantages of routinely collected administrative data for generating population level statistics:

1. Data quality: Administrative data may not be collected with the same level of rigor or accuracy as a census, and may contain errors or missing data.
2. Representativeness: Administrative data may not be representative of the entire population, as certain groups may be underrepresented or excluded from administrative records (e.g. homeless individuals, illegal immigrants, etc.).
3. Privacy concerns: The use of administrative data for population level analysis raises privacy concerns, as individuals' personal information may be used without their consent or knowledge.
4. Lack of detail: Administrative data may not provide the same level of detail as a census, and may not capture certain aspects of the population that are important for population level analysis (e.g. socio-economic status, cultural background, etc.).

**Question 6**

a)

Occupational diversity, as people in developing countries are more likely to work in a variety of occupations, including agriculture, manual labour, and informal jobs, whereas people in developed countries are more likely to work in specialised and highly skilled jobs.

Job security, as people in developing countries are more likely to work in insecure and poorly paid jobs, with limited benefits and protections, whereas people in developed countries are more likely to work in jobs with secure contracts, benefits, and protections.

b)

Occupational health surveillance systems, as these systems gather and analyse data on the health of workers, including information on workplace exposure to hazardous substances and the incidence of work-related illnesses and injuries.

Workers' compensation records, as these records contain information on the nature and frequency of work-related illnesses and injuries, and can provide valuable information on the health risks associated with specific occupations or industries.

Employee health surveys, as these surveys gather information on the health status, behaviours, and work conditions of employees, and can provide valuable insights into the factors that influence health at work.

Medical and health records, ashese records contain information on the medical history, treatment, and health status of workers, and can provide valuable information on the impact of work on health.

Job exposure matrix (JEM), as this tool is used to quantify the exposure of workers to hazardous substances, and can provide valuable information on the health risks associated with specific occupations or industries.

c) The proportional mortality ratio (PMR) is a measure of occupational mortality, calculated as the ratio of deaths from a specific cause in a specific occupational group compared to the expected number of deaths in a comparison population. PMR is used to identify occupations or industries that are associated with increased risk of death from a specific cause, and can be used to guide interventions to improve occupational health and safety. For example, a PMR of 1.5 would indicate that the observed number of deaths from a specific cause in a particular occupational group is 50% higher than the expected number of deaths in a comparison population.

Questions 7 and 8 removed as 10 mark questions

Question 9

a) Power and authority are related but distinct concepts. Power refers to the ability to influence others and make things happen, while authority refers to the legitimate right to make decisions and enforce rules. Power may be based on force, persuasion, or expertise, while authority is typically based on formal rules and institutional structures.

For example, a CEO may have the power to make decisions about the direction of an NHS trust, but their authority is based on their position within the organisational hierarchy and the rules and regulations that govern the trust.

b) In the context of an explosion in a large chemical factory in England, the roles of the police and fire emergency services would typically be to maintain public safety and control the spread of the fire, while the role of public health professionals would be to assess and manage the potential health impacts of the explosion, including the exposure of workers and the local population to hazardous chemicals.

For example, the police and fire services would be responsible for securing the scene and containing the fire, while public health professionals from the National Health Service (NHS) would be responsible for providing advice and support to those affected by the explosion, including workers and the local community.

c) The sources of power and authority for each profession in this emergency context would vary. The police and fire services would have the power and authority to enforce the law and maintain public order, while public health professionals would have the power to make decisions and provide advice based on their expertise and knowledge of the health impacts of chemical exposure.

In England, the police and fire services would have their power and authority derived from the law, and would be responsible to the government and the public, while public health professionals would have their power and authority derived from their professional training and expertise, and would be responsible to the NHS and the public.

Question 10 removed as 10 mark question

[**January 2015: questions and mark scheme**](https://www.fph.org.uk/media/1215/examiners-comments-and-key-points-june-15.pdf)

**Question 1**

a) A systematic review is a type of research methodology that aims to systematically and thoroughly identify, select, appraise, and synthesise all relevant evidence from multiple sources on a specific research question. Systematic reviews are widely recognised as the highest level of evidence in the hierarchy of evidence-based medicine.

One strength of the systematic review approach is that it provides a comprehensive and thorough examination of the available evidence, reducing the risk of bias and increasing the reliability and validity of the results. This can be especially useful in complex or controversial areas where there may be a large number of conflicting studies.

One limitation of the systematic review approach is that it can be time-consuming and resource-intensive, especially if a large number of studies need to be screened and appraised. Additionally, there may be publication bias and selective reporting, as studies with negative or inconclusive results may not be published, leading to an overestimation of the treatment effects.

b) Heterogeneity refers to the variability or differences between the studies included in a systematic review. It can refer to differences in study design, population, intervention, outcome measures, or other factors. Heterogeneity can impact the validity and reliability of the results of a systematic review, as it may indicate that the studies are not comparable or that the findings cannot be combined.

To assess heterogeneity, several methods can be used, including visual inspection of forest plots, chi-squared tests, I2 statistics, or meta-regression analysis. The choice of method will depend on the nature of the studies and the research question being investigated.

c) A funnel plot is a graphical representation of the distribution of study results that is used to detect publication bias in systematic reviews. The plot displays the effect size of each study on the vertical axis and the precision (inverse of the standard error) of each study on the horizontal axis. Ideally, the plot should be symmetrical, with small studies and large studies showing a similar range of effect sizes. If there is publication bias, then smaller studies with negative or inconclusive results may be missing, leading to a skewed or asymmetrical plot.

To interpret a funnel plot, it is important to look for signs of asymmetry, such as a funnel shape that is wider on one side than the other. If publication bias is suspected, the results of the systematic review should be interpreted with caution, as the true effect size may be different from the reported effect size. Additionally, other methods, such as regression analysis or trim-and-fill methods, can be used to adjust for publication bias and provide a more accurate estimate of the treatment effect.

Question 2

a) For qualitative research to understand why the physical activity campaign may not have been as successful as expected, several study methods can be employed. One method is conducting focus groups, which bring together a small group of children, parents, and teachers to discuss their experiences and opinions about the physical activity campaign. Another method is conducting in-depth interviews with individuals, to gain a deeper understanding of their perspectives and experiences. Observational studies, where children participating in physical activities are observed in different settings, can provide information on their physical activity behaviours and potential barriers to participation. Document analysis, reviewing relevant documents such as campaign materials and school policies, can give insight into the design and implementation of the campaign. Finally, online surveys can be used to collect data from a large sample of children, parents, and teachers, to gather information about their attitudes, behaviours, and experiences related to the physical activity campaign.
b) Qualitative research has both strengths and weaknesses in this situation. A strength is that it provides in-depth, rich information, allowing for a deeper understanding of complex phenomena and experiences. Additionally, qualitative research often involves participants in the research process, which can increase the validity and credibility of the findings. It also offers flexibility, allowing for the exploration of new and unexpected findings. However, a weakness is that qualitative research has limited generalisability, as the sample size is typically smaller than in quantitative research. The findings are also subjective, relying on the interpretations and perspectives of the researcher and participants, which can lead to bias and inconsistencies in the findings. Additionally, qualitative research is often time and resource intensive, requiring more time and resources to collect and analyse data compared to quantitative research methods.

Question 3 – removed due to 10 mark question

Question 4

a)

Genital Chlamydia infection is a highly prevalent sexually transmitted infection (STI) in England. According to the most recent data from Public Health England, Chlamydia is the most commonly diagnosed STI in the country, with an estimated 200,000 new cases reported each year. The highest rates of infection are seen among young people aged 15-24 years, and in certain ethnic minority populations. Additionally, Chlamydia is often asymptomatic, meaning that many infected individuals are unaware that they have the infection, and may unknowingly spread the infection to their sexual partners.

b) There are several population strategies that can be employed to reduce the disease burden of genital Chlamydia infection in England.

Increased testing involves encouraging individuals to get tested for Chlamydia on a regular basis, especially those who are sexually active, can help identify and treat infected individuals, reducing the spread of the infection.

Education and awareness involves providing education and awareness campaigns to target populations with high rates of Chlamydia, such as young people and ethnic minority populations, can help increase knowledge about the infection and promote safer sexual behaviours.

Access to care and treatment involves ensuring that individuals have access to prompt and effective care and treatment for Chlamydia can reduce the burden of the infection and prevent long-term health consequences.

Partner notification and treatment involves encouraging infected individuals to inform their sexual partners and encouraging those partners to get tested and treated can break the cycle of transmission and reduce the spread of the infection.

Condom use promotion - promoting consistent and correct condom use can help prevent the transmission of Chlamydia and other STIs.

Targeted interventions involves developing and implementing targeted interventions, such as screening programs and community outreach initiatives, in populations with high rates of Chlamydia can help reduce the burden of the infection and promote health equity.

Implementing these strategies in a comprehensive and coordinated manner can help reduce the burden of genital Chlamydia infection in England and improve sexual health outcomes for individuals and communities.

Question 5

a) The main causes of population expansion are as follows.

Increased life expectancy.

Advances in medicine and technology have led to an increase in life expectancy, allowing individuals to live longer and contributing to population growth.

Improved health care.

Improved access to health care and medical treatment has reduced mortality rates and increased the survival rates of individuals, leading to population growth.

Fertility rates.

High fertility rates, particularly in developing countries, have contributed to population growth. Cultural, economic, and religious factors can influence fertility rates and contribute to population growth.

Migration.

The movement of individuals and families from one geographic location to another, either within a country or across national borders, can contribute to population growth.

Natural increase.

The difference between the number of births and the number of deaths in a population can also contribute to population growth, particularly in countries with high birth rates.

b) Three policy approaches to reduce the rate of population growth include:

Family planning and reproductive health services.

Providing access to family planning and reproductive health services, including contraception and education, can help individuals and couples make informed decisions about their fertility and plan their families. This can help reduce the rate of population growth.

Education and empowerment of women.

Improving access to education and promoting gender equality can empower women and improve their health outcomes. Empowered women are more likely to have smaller families and engage in healthier behaviours, which can reduce population growth.

Sustainable development.

Promoting sustainable development and reducing poverty can improve health outcomes, reduce mortality rates, and reduce the incentives for families to have large families. This can help reduce population growth in the long-term.

The public health impact of these policies can be significant. Family planning and reproductive health services can improve maternal and child health, reduce maternal mortality, and prevent unintended pregnancies. Education and empowerment of women can improve health outcomes, reduce poverty, and improve gender equality.

Sustainable development can improve health outcomes, reduce poverty, and promote economic growth and stability. Implementing these policies at a national level can help reduce the rate of population growth and improve public health outcomes for individuals and communities.

Question 6

a) The crude mortality rate is a basic measure of the number of deaths in a population over a specified period of time. It is calculated as the number of deaths in a population divided by the total population, multiplied by a factor to reflect the size of the population. The crude mortality rate does not take into account differences in age distribution between populations, and so it is not a good indicator of differences in health status between populations.

The neonatal mortality rate is the number of deaths of infants less than 28 days of age per 1,000 live births. It is a measure of the health and survival of newborns and provides important information about the health and well-being of mothers and newborns.

For England, data sources used in the calculation of crude mortality rate and neonatal mortality rate include the Office for National Statistics (ONS) and the National Health Service (NHS) Digital. The ONS collects data on deaths and publishes annual mortality statistics, including information on cause of death and demographic information. The NHS Digital collects data on live births, stillbirths, and infant deaths and publishes annual statistics on neonatal and infant mortality.

b) Directly Standardised Mortality Rates (DSMR) are used to compare the mortality rates between populations with different age structures. This measure takes into account differences in age distribution between populations and provides a more accurate picture of differences in health status between populations. The DSMR is calculated by applying age-specific mortality rates from a standard population to a population of interest and summing the results.

Standardised Mortality Ratios (SMR) are used to compare the mortality rates of a population with a reference population. The SMR is calculated by dividing the observed number of deaths in a population by the expected number of deaths, based on the age-specific mortality rates of a reference population. The SMR is expressed as a ratio, with values greater than 1 indicating higher mortality rates and values less than 1 indicating lower mortality rates.

DSMR would be used when comparing mortality rates between populations with different age structures, while SMR would be used when comparing the mortality rates of a population with a reference population. Both measures are useful tools for understanding and comparing differences in mortality rates between populations and for identifying populations with increased health risks.

Question 7 and 8 – removed as 10 mark question

Question 9 – removed as 10 mark question

Question 10

a) In the context of healthcare, risk refers to the likelihood or probability of harm or negative outcomes occurring as a result of a particular intervention, treatment, or decision. This can include harm to patients, staff, or other stakeholders, as well as financial losses, operational disruptions, and other negative consequences. Understanding and managing risk is a critical part of providing safe, effective, and efficient healthcare.

b) To measure the level or scale of risk in healthcare, a framework is needed that considers multiple factors and dimensions. One commonly used framework is the Risk Management Matrix, which categorises risks based on their likelihood and potential impact. Another framework is the Healthcare Risk Control (HRC) Pyramid, which includes a hierarchical approach to risk management, starting with the identification and assessment of risks, followed by the implementation of controls to minimise or mitigate the risks, and ending with ongoing monitoring and evaluation to ensure the effectiveness of the risk management measures.

c) Healthcare organisations can minimise risk through the use of a comprehensive risk management strategy. This may include the following steps.

The first step in minimising risk is to identify and assess potential risks. This can be done through a variety of methods, including review of data and trends, conducting audits and assessments, and soliciting feedback from patients, staff, and other stakeholders.

Once risks have been identified and assessed, the next step is to develop and implement controls and mitigation measures to minimise or eliminate the risks. This may include changes to policies, procedures, and protocols, training and education programs, and the use of technology and other tools to improve safety and quality.

Ongoing monitoring and evaluation is critical to ensure that risk management measures are effective and that risks are continuously being identified and addressed. This may include ongoing review of data and trends, regular assessments and audits, and ongoing feedback from patients, staff, and other stakeholders.

Effective communication and collaboration among all stakeholders, including patients, staff, and healthcare organisations, is important for minimising risk. This may include regular engagement and consultation, development of effective communication and feedback channels, and the creation of partnerships and collaborations to share information, knowledge, and best practices.

By following these steps and incorporating risk management into the core operations of healthcare organisations, the risk of harm and negative outcomes can be minimised, leading to safer, more effective, and more efficient healthcare.

January 2016 – [questions and mark scheme](https://www.fph.org.uk/media/1213/questions-key-points-and-comments-for-web_january-2016.pdf)

Question 1

a) Four possible epidemiological explanations for the finding between the number of places licensed to sell alcohol in city centres and violent crime rates are:

The temporal association between the number of places licensed to sell alcohol and the violent crime rates may indicate a causal relationship. This means that the increase in the number of alcohol-selling establishments may be a direct cause of the increase in violent crime rates.

There may be other factors that are associated with both the number of alcohol-selling establishments and the violent crime rates. These confounding factors could be responsible for the observed relationship between the two variables, rather than the number of alcohol-selling establishments itself.

The spatial association between the number of alcohol-selling establishments and the violent crime rates may indicate that the concentration of alcohol-selling establishments in certain areas may be associated with higher violent crime rates.

It is also possible that the relationship between the number of alcohol-selling establishments and violent crime rates may be reversed. This means that the increase in violent crime rates may lead to an increase in the number of alcohol-selling establishments in the city centre.

b) In conducting a similar study in a different area, I would collect the following three variables in addition to the number of places licensed to sell alcohol and violent crime rates:

Demographic characteristics, such as age, gender, and socio-economic status, can influence both the number of alcohol-selling establishments and the violent crime rates. Thus, collecting data on demographic characteristics would help to control for potential confounding factors.

Law enforcement presence, such as the number of police officers and the frequency of patrols, may influence both the number of alcohol-selling establishments and the violent crime rates. Thus, collecting data on law enforcement presence would help to control for potential confounding factors.

Neighbourhood characteristics, such as the level of poverty, availability of public transportation, and availability of public spaces, may influence both the number of alcohol-selling establishments and the violent crime rates. Thus, collecting data on neighbourhood characteristics would help to control for potential confounding factors.

Question 2

a)

The increase in attendance rates could be due to an increase in the population of the area served by the hospital, leading to a greater demand for emergency services.

The increased attendance rates could be due to increased public awareness of the availability of emergency services, leading to more people seeking care.

The increase in attendance rates could be due to changes in health care delivery, such as an increase in the number of walk-in clinics or changes in the availability of primary care services, leading to more patients seeking care in the emergency department.

b)

The first step is to plan the evaluation, including setting clear goals and objectives, determining what data to collect, and identifying the methods to be used.

The second step is to collect data, which may include surveys, interviews, and/or observations, to determine the impact of the intervention.

The third step is to analyse the data collected, including summarising the results and interpreting the findings.

The fourth step is to report the results of the evaluation, including presenting the findings to key stakeholders and making recommendations for improvement.

The final step is to implement the recommendations for improvement, including making changes to the intervention and monitoring the results over time.

c) Three measures that I would use to evaluate the effectiveness of the "urgent care unit" are:

Patient satisfaction. It is an important measure of the effectiveness of a health service intervention. I would use surveys or interviews to assess patients' experiences with the urgent care unit, including the quality of care they received and the time they spent in the unit.

Clinical outcomes, such as the number of patients who are treated and released without being admitted to the hospital, are an important measure of the effectiveness of a health service intervention. I would use medical records and other data sources to track these outcomes and compare them with those of the emergency department.

Cost-effectiveness is an important measure of the effectiveness of a health service intervention. I would use data on the costs associated with the urgent care unit and the emergency department, including staffing costs, supplies, and equipment, to assess the cost-effectiveness of the urgent care unit.

Questions 3 and 4 – removed due to 10 mark questions

Question 5

a) To calculate life expectancy, you would require information on the mortality rate of a population, which can be obtained from a life table that tracks the number of deaths and survivors in a population over a given period of time. Other factors that could be useful to calculate life expectancy include data on age-specific death rates, birth rates, and migration patterns.

b) To calculate healthy life expectancy, you would need additional information on the health status and health behaviours of a population, such as data on risk factors for chronic diseases, access to health care, and health-promoting behaviours. This information would help to estimate the years lived in good health and the years lived with disability or disease.

Life expectancy and healthy life expectancy are used to describe the health status of a population and how this changes over time. Life expectancy provides an estimate of the average number of years a person can expect to live, while healthy life expectancy provides an estimate of the number of years a person can expect to live in good health. Both measures are important indicators of population health, as they provide information on the overall mortality and morbidity patterns in a population.

By tracking changes in life expectancy and healthy life expectancy over time, we can observe trends in population health and identify areas for improvement. For example, if life expectancy is increasing, it may indicate that the population is experiencing improved health and longevity, while a decline in healthy life expectancy may indicate an increase in the burden of chronic disease and disability. Monitoring these measures can help policymakers to identify and address public health issues and prioritise resources to improve population health.

Question 6

a) In England, there are several sources of routine data that can be used to measure mental health and mental wellbeing:

National Surveys - the Adult Psychiatric Morbidity Survey (APMS) and the Health Survey for England (HSE) are national surveys that provide information on the prevalence of mental disorders and mental health in the population. These surveys use standardised tools to assess the presence and severity of mental health conditions and gather information on mental health status, access to care, and other related issues.

Primary Care Data - GP practices in England routinely collect information on patient consultations, including data on diagnoses, treatments, and referrals. This data provides insight into the prevalence and treatment of mental health conditions in the primary care setting.

Hospital Episode Statistics (HES) - HES collects information on all admissions to National Health Service (NHS) hospitals in England. This data provides information on the hospital treatment of mental health conditions, including information on diagnosis, length of stay, and treatments received.

Mortality Data - the Office for National Statistics (ONS) collects information on all deaths in England, including deaths that are related to mental health conditions. This data provides information on the impact of mental health conditions on mortality and can be used to monitor trends and disparities in mental health outcomes over time.

(b) Despite the availability of these data sources, there are several limitations when using them to measure mental health and wellbeing in England:

Underdiagnosis and Undersampling - mental health conditions are often underdiagnosed, particularly in primary care settings, leading to a lack of accurate information on their prevalence and impact. Similarly, populations that are at higher risk for mental health conditions, such as ethnic minority populations and low-income groups, may be underrepresented in national surveys, leading to an underestimation of the true burden of mental health problems.

Stigma and Discrimination - the stigma and discrimination associated with mental health conditions can lead to underreporting and reluctance to seek treatment. This can result in a lack of accurate information on the prevalence and impact of mental health conditions.

Data Quality - the quality of mental health data collected in routine settings can vary, particularly in terms of its accuracy, completeness, and consistency. This can lead to limitations in the ability to use the data to measure mental health outcomes and monitor trends over time.

Data Linkage - linking data across different sources, such as primary care and hospital data, can be challenging, particularly in terms of ensuring the accuracy and completeness of the data. This can limit the ability to obtain a comprehensive view of mental health outcomes and the impact of interventions.

Question 7 – removed as 10 mark question

Question 8 – removed as 10 mark question

Question 9

a) The difference between strategy and policy in the context of tobacco control refers to the level of detail and the scope of the action that is being proposed.

A policy is a formal, written statement of the rules, principles, and guidelines that are to be followed in a particular area. Policies provide a framework for decision making and serve as a guide for those responsible for implementing them. In the context of tobacco control, a policy might outline the overall goals and objectives of the government or organisation in reducing tobacco use, such as increasing taxes on tobacco products, regulating advertising and promotion, or providing support for smokers to quit.

A strategy, on the other hand, is a plan of action that outlines the specific steps that will be taken to achieve a particular goal. Strategies provide a roadmap for achieving the policy objectives and are often more detailed and specific. For example, a strategy to reduce tobacco use might include specific measures such as implementing smoke-free laws, launching mass media campaigns to promote quitting, or increasing access to smoking cessation treatments.

(b) To improve tobacco control, I would choose to use the Framework Convention on Tobacco Control (FCTC), which is a treaty established by the World Health Organisation (WHO) to provide a comprehensive framework for global tobacco control.

The FCTC provides a comprehensive set of measures to reduce the demand for tobacco, including measures related to tobacco taxation, advertising, and promotion, as well as measures to support smokers to quit.

In order to improve tobacco control, the following actions could be taken:

- Raising taxes on tobacco products is one of the most effective ways to reduce tobacco use, particularly among youth and lower-income populations.
- Establishing smoke-free environments, such as restaurants, bars, and public places, can reduce exposure to second-hand smoke and create a supportive environment for smokers to quit.
- Restricting the advertising and promotion of tobacco products, particularly to youth, can reduce the impact of marketing on tobacco use and can help to counter the effects of marketing on smoking initiation.
- Providing access to effective smoking cessation treatments, such as nicotine replacement therapy and behavioural support, can increase the likelihood of smokers quitting and reducing their exposure to the harmful effects of tobacco.
- Launching mass media campaigns to raise awareness about the dangers of smoking and the benefits of quitting can help to motivate smokers to quit and can change social norms around tobacco use.
- Strengthening enforcement of tobacco control measures, such as restricting the sale of tobacco products to minors and ensuring that products are properly labelled, can increase the effectiveness of tobacco control policies.
- Regularly monitoring and evaluating the impact of tobacco control policies and strategies, and making adjustments as needed, can help to ensure that they are effective and achieving their intended goals.

Question 10

Question removed as 10 mark question

[June 2016: Questions and mark scheme](https://www.fph.org.uk/media/1212/part-a-mfph-june-16-questions-key-points-and-comments.pdf)

**Question 1**

a) Clustered randomised trials are appropriate in certain cases because they reflect the natural unit of intervention delivery and reduce contamination.

Natural unit of intervention delivery. Clustered randomised trials are appropriate when the intervention is delivered to groups rather than individuals. For example, in a school-based intervention to prevent obesity, the natural unit of intervention delivery is the school rather than the individual student.

Contamination. Clustered randomised trials can be used to reduce contamination between intervention and control groups. For example, in a community-based intervention to improve physical activity, contamination could occur if individuals in the control group adopt the intervention behaviours after observing the intervention group.

b) Two disadvantages of cluster randomised trials are: 1) higher cost and 2) design complexity.

Higher cost - cluster randomised trials are more expensive than individual randomised trials due to the need for larger sample sizes to account for the clustering effect.

Design complexity – cluster randomised trials are more complex to design and implement than individual randomised trials. For example, they require adequate randomisation of clusters and may need to consider stratification or blocking to ensure balance between intervention and control groups.

c) In cluster randomised trials, the sample size estimation needs to take into account the intra-cluster correlation, which is the correlation between individuals within a cluster. This increases the variance of the treatment effect estimate, and therefore a larger sample size is needed to achieve the same level of precision as in an individual randomised trial. To estimate the sample size for a cluster randomised trial, the design effect, which is the ratio of the variance of the treatment effect estimate in a cluster randomised trial to the variance of the estimate in an individual randomised trial, must be taken into account. Additionally, the cluster size, the intracluster correlation coefficient, and the desired level of precision must be considered in sample size calculation.

Question 2

a)

1. Knowledge and training of healthcare professionals: Variation may arise due to differences in the knowledge and training of healthcare professionals in recognising and managing diabetic ketoacidosis in different general practices.
2. Patient-provider communication: Variation may occur due to differences in patient-provider communication regarding symptoms and management of diabetic ketoacidosis in different general practices.
3. Availability of resources: Variation may be due to differences in the availability of resources, such as glucose meters, insulin, and testing supplies, in different general practices.
4. Disease management protocols: Variation may result from differences in the use of disease management protocols for diabetic ketoacidosis in different general practices.

b)	Key features of a clinical audit of the management of diabetic ketoacidosis in the community, such as in general practice, require a clear definition of the population being studied. For example, the population could be defined as all patients with diabetic ketoacidosis managed in general practice. It is also important to establish audit criteria to assess the quality of care provided. This could include assessment of the time to treatment initiation, use of standard protocols, and patient outcomes.

In order to assess compliance with these criteria, data should be collected systematically and consistently. This can be done through chart reviews, patient interviews, and/or questionnaires. Once the data has been collected, it should be analysed and interpreted to identify areas of improvement and opportunities for change. The results of the audit should be reported back to the healthcare professionals and organisations involved, with recommendations for improvement where necessary. Finally, the recommendations for improvement should be implemented and monitored to ensure that changes are sustained.

Question 3

a) In the UK, seasonal influenza is a common and highly contagious respiratory illness caused by the influenza virus. It can lead to serious complications, particularly in vulnerable populations such as the elderly, young children, pregnant women, and people with chronic health conditions. Each year, the epidemiology of seasonal influenza is influenced by the circulation of new strains of the virus, population immunity, and the effectiveness of the annual influenza vaccine.

b) To organise an effective local influenza immunisation program and ensure high uptake in all relevant groups in the UK, the following steps could be taken:

Partner with local healthcare providers. Partnering with local healthcare providers, such as GP surgeries, community health clinics, and hospitals, can help to reach a large portion of the target population and ensure that they are offered the vaccine.

Increase awareness and education. Increasing awareness and education about the importance of influenza vaccination can help to dispel myths and encourage uptake. This can be achieved through a variety of methods, such as public health campaigns, community events, and educational materials distributed through healthcare providers.

Offer convenient vaccine access. Offering convenient access to the vaccine, such as through on-site clinics or home visits, can increase uptake among those who may face barriers to accessing healthcare services.

Target high-risk populations. Targeting high-risk populations, such as the elderly and people with chronic health conditions, can help to ensure that those who are most vulnerable to the complications of influenza receive the vaccine.

Monitor and evaluate the program. Monitoring and evaluating the program can help to identify areas of success and areas that need improvement. This can be achieved through tracking vaccine uptake, conducting surveys of program participants, and analysing data on influenza-related hospitalisations and deaths.

Sustain the program. Sustaining the program over time can help to ensure that high uptake is maintained and that the program continues to have a positive impact on public health. This can be achieved through ongoing public health campaigns, partnerships with healthcare providers, and ensuring that the vaccine remains accessible and convenient for all target populations.

Question 4 – removed due to 100% mark for single question

Question 5

a) In England, the following types of routine data, including key characteristics of the population, would be important to include in an electronic information system to support the planning of healthcare services:

- Demographic data, such as age, gender, ethnicity, and socioeconomic status.
- Clinical data, such as previous medical history, current medications, and conditions.
- Epidemiological data, such as the incidence and prevalence of certain diseases and conditions in the population.
- Health behaviours data, such as smoking, alcohol consumption, and physical activity levels.
- Health outcomes data, such as mortality and morbidity rates, hospitalisation rates, and readmission rates.
- Resource utilisation data, such as the number of outpatient visits, hospital admissions, and length of stay.
- Geographical data, such as the location and accessibility of healthcare facilities and services.
- Socioeconomic data, such as income, education, and employment status.

b) In an electronic information system to support service planning, there are several key capabilities that are important to have. Firstly, the system should allow for data visualisation. This means that the data can be viewed and analysed in a visual format, such as charts, graphs, and maps, to help identify trends and patterns in the population's health status and resource utilisation. This can provide valuable insights into the current situation and help guide decision making about service planning.

Another important aspect of the system is data integration. This means that data from different sources can be combined and analysed to get a more complete picture of the population's health status and resource utilisation. For example, data from hospitals, general practitioners, and public health agencies can be combined to provide a more comprehensive view of the population's health status. This can help to identify areas where collaboration and coordination in service planning is needed.

Data sharing is also an important aspect of the system. This allows for the data to be shared between different organisations and stakeholders, such as hospitals, general practitioners, and public health agencies. This can help to support collaboration and coordination in service planning, and can also help to ensure that all stakeholders have access to the same information.

In addition, the system should allow for data analysis. This means that statistical analysis can be performed on the data to identify trends, patterns, and relationships between variables. This information can then be used to inform decision making about service planning, and to identify areas where improvements are needed.

Another important aspect of the system is data forecasting. This means that historical data and trends can be used to predict future health status and resource utilisation. This can help to plan for future service needs, and ensure that resources are allocated where they are needed most.

Finally, performance monitoring is also important in an electronic information system to support service planning. This means that the performance of healthcare services can be monitored against key performance indicators, such as patient satisfaction and clinical outcomes. This information can then be used to make improvements where necessary and ensure that healthcare services are providing the best possible care to patients.

**Question 6**

a) To calculate a forecast of the number of people with dementia in a local area in England in 10 years' time, one could use a mathematical model such as the age-period-cohort (APC) model. This model takes into account factors such as age, time period, and cohort effects to predict the number of cases of dementia in a given population. The model would require data on the current number of people with dementia in the local area, as well as data on demographic and epidemiological trends such as population growth, aging, and changes in risk factors for dementia.

b) When calculating a forecast using a mathematical model such as the APC model, it is important to consider key assumptions. One key assumption is that the demographic and epidemiological trends that have existed in the past will continue into the future. This includes assumptions about population growth, aging, and changes in risk factors for dementia. Another key assumption is that there will be no major breakthroughs in the prevention or treatment of dementia that would significantly impact the number of cases.

It is also important to consider other factors that could affect the accuracy of the forecast. For example, changes in the availability or quality of healthcare services in the local area could impact the number of cases of dementia. Additionally, changes in social and lifestyle factors, such as increased physical activity and improved diet, could also impact the number of cases of dementia.

c) Changes in dementia care could have a significant impact on the mathematical modelling of a 10-year forecast. For example, if there are major breakthroughs in the prevention or treatment of dementia, this could lead to a reduction in the number of cases. Conversely, if there are cuts to healthcare services or a reduction in the availability of support for people with dementia, this could lead to an increase in the number of cases.

It is also important to consider the impact of changes in demographics and population structure. For example, if the population in the local area is aging rapidly, this could lead to an increase in the number of cases of dementia. On the other hand, if the population is becoming younger, this could lead to a decrease in the number of cases.

Overall, it is important to consider a range of possible scenarios when assessing the impact of changes in dementia care on a 10-year forecast. This could include scenarios such as breakthroughs in prevention and treatment, changes in healthcare services, and changes in demographics and population structure. By considering a range of scenarios, it is possible to develop a more accurate and reliable forecast of the number of people with dementia in a local area in England in 10 years' time.

Question 7

a) Uptake of cervical screening in England can vary significantly between different groups in society. From a sociological perspective, differences in uptake can be linked to a range of factors, including socio-economic status, ethnicity, and education. Women from lower socio-economic groups, for example, may be less likely to attend cervical screening due to a range of barriers, such as poverty, low levels of education, and limited access to healthcare services.

Differences in uptake can also be linked to cultural and religious beliefs. For example, some women from ethnic minority groups may feel uncomfortable undergoing a cervical screening due to cultural or religious reasons or may be reluctant to attend due to fears about confidentiality and privacy. Women who have limited access to information about the benefits of cervical screening, or who lack understanding about the importance of the test, may also be less likely to attend.

b) To reduce inequalities in uptake of cervical screening between different groups in society, it is important to address the underlying social, cultural, and economic factors that contribute to these differences. This could include targeted campaigns and outreach initiatives to raise awareness about the importance of cervical screening, as well as the provision of clear and accessible information about the test and the benefits it provides.

It is also important to consider the role of healthcare providers in reducing inequalities in uptake. Healthcare providers can play a key role in ensuring that women from different groups in society have equal access to screening services and that they are provided with the information and support they need to make an informed decision about whether to attend.

Other strategies to reduce inequalities in uptake could include improving access to healthcare services, particularly in areas with high levels of poverty or where there are high numbers of women from ethnic minority groups. Providing flexible appointment times, or offering screening services at community centres or other local venues, could also help to overcome barriers to attendance.

In conclusion, reducing inequalities in uptake of cervical screening requires a multi-faceted approach that addresses the underlying social, cultural, and economic factors that contribute to these differences. This could include targeted campaigns and outreach initiatives, improved access to healthcare services, and the provision of clear and accessible information about the benefits of cervical screening.

Question 8

Removed due to being a 10 mark question

Question 9

a) The use of targets as an improvement tool in health services in England has several disadvantages. One disadvantage is the possibility of unintended consequences. This can occur when the implementation of targets leads to changes in practice that are not in the best interest of patients or negatively impact patient safety. For example, a target to reduce waiting times for outpatient appointments could result in clinicians rushing through consultations, leading to decreased patient satisfaction and suboptimal care.

Another disadvantage is the narrow focus of targets. They can often be too narrowly focused, neglecting other important aspects of patient care. For example, a target to increase the number of cancer patients receiving a certain type of treatment could result in a neglect of other important aspects of care, such as symptom management or patient support.

The use of targets can also result in a lack of accountability, with individuals or organisations being held responsible for achieving the target but not necessarily for the impact of their actions on patients or the wider health system. This can be problematic as it can lead to actions being taken that are not in the best interest of patients.

Targets can also discourage innovation and creativity, as individuals and organisations may be too focused on achieving the target to consider alternative approaches or new ideas. This can limit the ability of the health service to improve and evolve, leading to stagnation.

Finally, the use of targets relies on accurate and reliable data. However, this is not always the case and inaccurate or incomplete data can lead to targets being misaligned with actual needs or to incorrect conclusions about the effectiveness of health services. This can result in suboptimal care for patients and the waste of resources.

b) A good target can serve as an effective improvement tool for health services in England if it possesses certain features. Firstly, a good target should be relevant to the specific health service in question and aligned with the overall goals and objectives of the service. For example, if the health service aims to improve patient satisfaction, a relevant target might be to increase patient satisfaction scores by a certain percentage within a specific time frame.

Secondly, a good target should be measurable, with clear metrics and data that can be used to assess progress and determine whether the target has been met. For example, in the above scenario, patient satisfaction scores could be measured using a standardised survey.

Thirdly, a good target should have a clear timeline for achievement, with specific dates or deadlines for completion. This helps to ensure that the health service is focused on achieving the target in a timely and efficient manner.

Fourthly, a good target should take into account the needs and priorities of patients, being patient-centred and focused on delivering the best possible care. For example, a target to reduce waiting times for appointments might take into account patient feedback on preferred appointment times.

Finally, a good target should be realistic, taking into account the resources and capabilities of the health service. This helps to ensure that the target is achievable, and that the health service is not set up for failure by setting unrealistic expectations. For example, a target to reduce waiting times for appointments may consider staffing levels, available technology, and patient volumes.

Question 10

a) One relevant motivational theory in public health practice in England is Self-Determination Theory (SDT).

Self-Determination Theory proposes that motivation is driven by three innate psychological needs: autonomy, competence, and relatedness. Autonomy refers to the need for control over one's own life and decisions. Competence refers to the need to feel effective and capable in one's actions. Relatedness refers to the need for connection and support from others.

According to SDT, when these three needs are met, individuals are more likely to experience intrinsic motivation and engage in activities with greater engagement, enthusiasm, and persistence. On the other hand, when these needs are thwarted, individuals may experience reduced motivation and engagement.

b) As a leader of a public health team during a period of major organisational change, I would use Self-Determination Theory to support my team through this period of uncertainty. By considering the needs of autonomy, competence, and relatedness, I can help ensure that my team members feel in control of their own work, confident in their abilities, and connected to one another and the organisation.

To support autonomy, I would encourage team members to take ownership of their work and make decisions for themselves, while also providing clear guidelines and resources for decision-making. To support competence, I would provide opportunities for training and professional development, and offer positive feedback and recognition for their efforts. To support relatedness, I would encourage team members to work together and build strong relationships with one another, while also promoting a supportive and inclusive work environment.

By addressing these three needs, I believe I can help my team feel more motivated and engaged in their work, even during a time of change. This, in turn, can help support the success of the organisational changes and ultimately lead to improved public health outcomes.

[**January 2017 questions and mark scheme**](https://www.fph.org.uk/media/1211/key-points-examiner-comments-jan17-final.pdf)

**Question 1**

a) The key stages of a systematic review are:
1. Formulating the research question and defining inclusion criteria
2. Searching for relevant studies
3. Selecting studies for inclusion in the review
4. Assessing the quality of the studies
5. Extracting data from the studies
6. Synthesising the data from the studies
7. Interpreting the results and drawing conclusions
8. Reporting the results in a systematic and transparent manner.
b) I2 of 50% in this systematic review indicates moderate heterogeneity among the studies included in the analysis. Heterogeneity refers to the differences among the studies in terms of their methods, results, and populations studied. An I2 of 50% suggests that half of the observed variability in results among the studies is due to true differences, and half is due to chance.
c) The possible reasons for the lack of clear effects of reducing saturated fat intake on total and cardiovascular mortality, as stated by the authors, could include:
1. Short duration of follow-up: The studies included in the systematic review may not have followed participants for a long enough period of time to detect significant differences in mortality outcomes.
2. Confounding factors: Other factors besides dietary fat intake, such as lifestyle, socio-economic status, and genetics, may have affected the results and diluted the effect of dietary changes on mortality.
3. Lack of adherence: Participants may have not adhered to the prescribed dietary changes, which would limit the impact of reducing saturated fat on mortality outcomes.
4. Insufficient sample size: The sample size of the included studies may have been too small to detect significant differences in mortality outcomes.
5. Other dietary factors: Changes in other aspects of the diet besides saturated fat intake, such as the intake of polyunsaturated fats or carbohydrates, may have masked the effect of reducing saturated fat on mortality.

**Question 2**

a) Key features of qualitative research are:

1. Emphasis on understanding the experiences, perspectives, and meanings of participants.
2. Use of unstructured or semi-structured data collection methods, such as interviews, focus groups, and observation.
3. Collection of rich, in-depth data through open-ended questions and follow-up probes.
4. Data analysis that often involves themes, categories, and patterns emerging from the data, rather than quantifying results.
5. Flexibility and adaptability in the research process, as the researcher can modify their approach as new information emerges.
6. Attention to the context and environment in which the research takes place.
7. A focus on the participants' perspectives and experiences, rather than objectively 'measuring' a phenomenon.

b) In a local community affected by Female Genital Mutilation (FGM), a relevant group could be women who have undergone FGM or are at risk of undergoing FGM. One appropriate qualitative method of data collection in this group could be in-depth interviews. In-depth interviews involve one-on-one conversations with participants, typically lasting between 30 minutes to an hour. This method is appropriate because:

1. It allows the researcher to explore complex experiences and attitudes in detail.
2. Participants are able to express their thoughts and feelings in their own words, rather than being limited by structured questions.
3. The researcher can follow-up with additional probes and questions, further exploring the participants' experiences and attitudes.
4. The method is less intimidating for participants, as it does not involve group dynamics that can be challenging for some individuals.
5. The method can be conducted in a private and confidential setting, reducing the risk of stigma or social pressure on participants.
6. The method can provide rich and in-depth data, allowing the researcher to understand the perspectives and experiences of the participants in a nuanced manner.

Question 3 – removed as 10 mark question

**Question 4**

a) In the United Kingdom, lung cancer is a significant public health issue, accounting for a large proportion of cancer-related deaths. Key epidemiological features of lung cancer in the UK include:

1. High incidence rate: Lung cancer is one of the most common cancers in the UK, with over 44,500 new cases diagnosed each year.
2. Age distribution: Lung cancer is more common in older people, with most cases occurring in those over the age of 60.
3. Gender disparity: Lung cancer is more common in men than women, with a male to female ratio of 2:1.
4. Smoking as a risk factor: Smoking is the main risk factor for lung cancer, responsible for around 85% of cases in the UK.
5. Variation in incidence by region: There is variation in the incidence of lung cancer across different regions of the UK, with higher rates in more deprived areas.

b) Healthcare services play a crucial role in the primary prevention of lung cancer by promoting healthy behaviours, screening for early detection, and providing interventions for those at high risk. Some of the ways healthcare services contribute to the primary prevention of lung cancer include:

1. Smoking cessation support: Healthcare services provide advice and support to help people quit smoking, the main preventable risk factor for lung cancer.
2. Screening programmes: Lung cancer screening programmes can detect the disease at an early stage, when it is more treatable.
3. Health promotion and education: Healthcare services provide information and education to the public about the risks of smoking and the benefits of healthy behaviours, such as regular exercise and a healthy diet.
4. Risk assessment and early detection: Healthcare services offer risk assessments and diagnostic tests to identify individuals who may be at higher risk of developing lung cancer, allowing for early detection and treatment.
5. Treatment and support: Healthcare services provide access to treatments, such as surgery, chemotherapy, and radiation therapy, as well as support for patients and their families.

**Question 5**

a)

Comprehensive data collection: The register should collect data on all cases of the disease in the population, not just those seen by healthcare providers.

Standardised data collection: The register should use a standardised data collection method to ensure that data is collected consistently across cases.

Data quality assurance: The register should have a system in place to ensure the accuracy and completeness of the data collected.

Linkage to other data sources: The register should be linked to other relevant data sources, such as hospital records and death certificates, to provide a complete picture of the disease.

Confidentiality: The register should ensure the confidentiality of patient information and comply with data protection laws.

b)

Data collection: Collecting data on all cases of the disease in a population can be challenging, as some cases may not be diagnosed or may not come to the attention of healthcare providers.

Data quality: Ensuring the accuracy and completeness of data can be difficult, especially if data is collected from multiple sources with different levels of accuracy.

Data protection: Ensuring the confidentiality of patient information and compliance with data protection laws can be challenging, particularly if the register holds sensitive information.

Funding: Establishing and maintaining a population-based disease register can be expensive, and ongoing funding may be difficult to secure.

c)

Disease incidence and prevalence: The register can provide information on the number of new cases of the disease and the number of people living with the disease in the population.

Disease distribution: The register can provide information on where cases of the disease are occurring, which can inform the planning of healthcare services in areas with higher disease burden.

Risk factors: The register can provide information on risk factors for the disease, which can inform the development of targeted prevention strategies.

Treatment and outcomes: The register can provide information on treatment patterns and outcomes, which can inform the development of more effective treatments and improve patient outcomes.

**Question 6**

a) Direct age-standardised death rates are calculated by dividing the number of deaths in a specific age group by the size of the corresponding population, multiplied by a standard population. In this case, the deaths of men aged 65 and over in each region are divided by the size of the older male population in each region, and then multiplied by a standard population (100,000) to give a death rate per 100,000 men.

b) Limitations of the direct age-standardised death rates in giving a picture of accidents in older people are:

- They only take into account the number of deaths and not the number of accidents.
- They may not account for differences in the size and age structure of the populations of different regions.
- They do not take into account the impact of access to healthcare services and the quality of care.
- They do not reflect the socio-economic status, lifestyle, and environmental factors that may contribute to differences in death rates between regions.

c) To monitor the health of older people regarding accidents, a public health practitioner may:

- Collect data on both accidents and fatalities using a combination of sources such as hospital records, mortality data, and emergency department visits.
- Track trends over time and compare the data to other populations to identify any disparities.
- Conduct community-based surveys to gather information about the attitudes, beliefs, and lifestyles of older people.
- Partner with local organisations and community groups to raise awareness about safe practices and promote injury prevention initiatives.

**Question 7**

a) The impact of migration on the health of migrants can be significant and can affect their physical, mental and social well-being. Migration can bring new stressors and challenges such as language barriers, cultural differences, poverty, discrimination, and social isolation, which can lead to poor health outcomes. Migrants may also face difficulty accessing healthcare services, especially those without proper documentation, leading to an increased risk of disease and decreased access to preventative care. In some cases, exposure to hazardous work or living conditions can also have a negative impact on their health. On the other hand, migration can also bring new opportunities and improved living conditions, which can have a positive impact on their health.

b) The impact of migration on the country to which they migrate can be both positive and negative. On the positive side, migrants bring diversity, skills, and new perspectives to the country, which can help boost the economy and cultural richness. However, migration can also put a strain on public services such as healthcare and education, as well as lead to social tensions, discrimination and increased poverty in certain communities. The integration of migrants into society can also be a challenge, which can lead to isolation, poor mental health, and the perpetuation of cultural stereotypes. To mitigate the negative effects of migration on society, it is important for the country to have well-designed policies and programmes that address the needs of migrants and promote their integration into the community.

**Question 8**

a) From a sociological perspective, there are several challenges and barriers that need to be overcome in order to enable patient and public involvement in health planning and policy development. One major challenge is the power imbalance between patients and healthcare providers, as patients may feel that their opinions and needs are not being heard or valued. Additionally, many patients may not feel confident or knowledgeable enough to participate in these types of activities, especially if they are marginalised or have limited access to information and resources. Other barriers may include language barriers, cultural differences, and limited opportunities for engagement and feedback.

b) An example of a patient and public involvement group could be a patient advisory council, which is composed of patients and members of the public who provide feedback and advice on various aspects of healthcare delivery and policy. The benefits of involvement in such a group include increased patient satisfaction, improved healthcare quality and outcomes, and greater transparency and accountability in the healthcare system. Patients and members of the public can provide valuable perspectives and experiences that help to inform decision-making and shape policies and programmes that better meet the needs of the community.

c) To ensure that patients and members of the public are enabled to participate fully in a patient and public involvement group, it is important to provide training and support, and to foster a culture of openness and collaboration. This could include providing training on public speaking, healthcare policy, and patient engagement, as well as providing access to resources and information. It may also be helpful to establish clear guidelines and processes for participation and decision-making, and to ensure that patients and members of the public have equal opportunities to contribute and have their voices heard. Other measures may include providing stipends or reimbursement for time and expenses incurred in participating, and regularly seeking and responding to feedback from group members to ensure that their needs and opinions are being heard and addressed.

**Question 9**

(a) To understand the individual and collective capacities of the group members, I would take the following actions. First, I would schedule individual meetings with each group member to get a clear understanding of their strengths, weaknesses, interests, and motivations. This would allow me to better understand how each member can contribute to the group's success. Secondly, I would encourage open communication among group members and foster an environment that promotes trust and collaboration. This would help to build relationships and ensure that everyone is comfortable sharing their thoughts and ideas. Finally, I would conduct group activities and assessments such as personality tests, strengths finder assessments, or team-building exercises to understand group dynamics and collective capacities. These activities would help to identify the group's strengths and weaknesses, as well as areas for improvement.

(b) I would apply Bruce Tuckman's theory of group development, also known as "Tuckman's stages of group development." According to this theory, groups go through five stages: forming, storming, norming, performing, and adjourning. During the forming stage, the group members are polite and cautious, trying to get to know each other and understand the task at hand. In the storming stage, there may be conflicts and disagreements as group members begin to express their opinions and compete for control. The norming stage involves resolving conflicts, establishing group norms, and developing a sense of cohesion. In the performing stage, the group is highly productive, working efficiently and effectively towards the common goal. Finally, in the adjourning stage, the group disbands once the task is completed. This theory provides a roadmap for understanding and managing the development of a group over time.

(c) I would use transformational leadership theory to develop this group. According to this theory, a transformational leader inspires and motivates followers to not only achieve the set goals but also strive for personal growth and development. I would lead by example and embody the values and vision of the group. This would help to establish trust and credibility with the group members. Additionally, I would communicate the purpose and vision of the group in a clear and inspiring manner, which would help to motivate and engage the group. I would also provide individualised consideration, recognising the unique needs and strengths of each group member and providing opportunities for growth and development. This would help to foster a sense of belonging and engagement among the group members. Finally, I would encourage innovation and risk-taking, creating a safe environment for members to express their ideas and experiment with new solutions. This would help to generate new and innovative solutions to the complex public health concern.

**Question 10**

a) For the delivery of the new obesity strategy, I would choose a Stakeholder Analysis as a management tool to help understand the current situation. This tool is appropriate and useful in this context as it helps to identify and assess the interests, influence, and potential impact of different stakeholders on the implementation of the obesity strategy. The strengths of Stakeholder Analysis include its ability to provide a comprehensive overview of the stakeholders involved and their potential impact, as well as its ability to facilitate effective communication and engagement with stakeholders. The weaknesses of Stakeholder Analysis include its subjective nature, as the results are dependent on the perspective and interpretation of the individual conducting the analysis, and its tendency to oversimplify complex stakeholder relationships.

b) To help implement the obesity strategy, I would use Kotter's 8-Step Change Model. This model is appropriate in this context as it provides a structured approach to managing change, which is critical for the successful implementation of a new obesity strategy. The eight steps of Kotter's model are: establish a sense of urgency, create a guiding coalition, develop a vision and strategy, communicate the change vision, empower others to act on the vision, create short-term wins, consolidate gains and produce more change, and anchor new approaches in the organisation's culture. The strengths of Kotter's model include its focus on leadership and communication, which are critical components of change management, and its attention to the importance of creating a sense of urgency and establishing a clear vision for change. The weaknesses of Kotter's model include its linearity, as real-life change is often more complex and non-linear, and its emphasis on top-down change, which may not always be feasible or effective in all contexts.
