## Supplementary material - example of unique insight for "ChatGPT sits the DFPH exam: large language model performance and potential to support public health learning"

**Example of what constitutes a unique insight**

The paper uses a modified definition of insight that builds on the work of [Kung 2023](https://www.tandfonline.com/doi/pdf/10.1207/s15430421tip4104_2?casa_token=3dZVomPjZmMAAAAA:_oI8SgWOh844xxftGqxsk7a2f9RduMamt8GpgmSU3aajorO61T9VmiPm5YhPRSb22EGsmwozO9T9).

An insight must meet the following three conditions:

- Nondefinitional: Does not simply define a term in the input question
- Nonobvious: Requires deduction or knowledge external to the question input
- Valid: Is in keeping with public health practice or numerically accurate; preserves directionality

Example

Q. What are the strengths and weaknesses of semi-structured interviews as a qualitative method for research on a weight management service?

A. Semi-structured interviews are one-on-one interviews used in qualitative research that follow a broad question guide, although interviewers may ask follow-up questions not on the guide to elicit data to support study aims. ***(not insight – simply defines input term).*** They can be used in weight management. **(*not insight – only uses internal question information).*** They are far weaker than quantitative studies as they do not make use of numerical empirical data. (***not insight – would not be recognised as valid***). However, in collecting perspectives of those who use the weight management service, barriers and facilitators to participants achieving a healthy weight could be identified, e.g. the attitudes and approaches of instructors (***1x instance of insight – meets all three criteria*.)**
